# Remote Covid Assessment in Primary Care (RECAP) risk prediction tool: derivation and real-world validation studies

**DOI:** 10.1101/2021.12.23.21268279

**Authors:** Ana Espinosa-Gonzalez, Denys Prociuk, Francesca Fiorentino, Christian Ramtale, Ella Mi, Emma Mi, Ben Glampson, Ana Luisa Neves, Cecilia Okusi, Laiba Hussain, Jack Macartney, Martina Brown, Ben Browne, Caroline Warren, Rachna Chowla, Jonty Heaversedge, Trisha Greenhalgh, Simon de Lusignan, Erik Mayer, Brendan Delaney

## Abstract

**Background:** Accurate assessment of COVID-19 severity in the community is essential for best patient care and efficient use of services and requires a risk prediction score that is COVID-19 specific and adequately validated in a community setting. Following a qualitative phase to identify signs, symptoms and risk factors, we sought to develop and validate two COVID-19-specific risk prediction scores RECAP-GP (without peripheral oxygen saturation (SpO2)) and RECAP-O2 (with SpO2).

**Methods:** Prospective cohort study using multivariable logistic regression for model development. Data on signs and symptoms (model predictors) were collected on community-based patients with suspected COVID-19 via primary care electronic health records systems and linked with secondary data on hospital admission (primary outcome) within 28 days of symptom onset. Data sources: RECAP-GP: Oxford-Royal College of General Practitioners Research and Surveillance Centre (RSC) primary care practices (development), Northwest London (NWL) primary care practices, NHS COVID-19 Clinical Assessment Service (CCAS) (validation). RECAP-O2: Doctaly Assist platform (development, and validation in subsequent sample). Estimated sample size was 2,880 per model.

**Findings:** Data were available from 8,311 individuals. Observations, such SpO2, were mostly missing in NWL, RSC, and CCAS data; however, SpO2 was available for around 70% of Doctaly patients. In the final predictive models, RECAP-GP included sex, age, degree of breathlessness, temperature symptoms, and presence of hypertension (Area Under the Curve (AUC): 0.802, Validation Negative Predictive Value (NPV) of ‘low risk’ 98.8%. RECAP-O2 included age, degree of breathlessness, fatigue, and SpO2 at rest (AUC: 0.843), Validation NPV of ‘low risk’ 99.4%.

**Interpretation:** Both RECAP models are a valid tool in the assessment of COVID-19 patients in the community. RECAP-GP can be used initially, without need for observations, to identify patients who require monitoring. If the patient is monitored at home and SpO2 is available, RECAP-O2 is useful to assess the need for further treatment escalation.

**Research in context panel:** *Evidence before the study:* This study was conceived during the first COVID-19 wave in the UK (March - April 2020), as members of the research team contributed to the development of national clinical guidelines for COVID-19 management in the community and to the Oxford COVID-19 rapid review to track signs and symptoms of COVID-19 internationally. The review was carried out according to Cochrane Collaboration standards for rapid reviews and identified systematic reviews and large-scale observational studies describing the signs and symptoms of COVID-19. Evidence gathered showed worsening of COVID-19 symptoms around the 7th day of disease and challenges in identifying patients with higher likelihood of severity to increase their monitoring. To this end, tools such NEWS2 have been used in the UK to assess COVID-19 patients in primary care, but they do not capture the characteristics of COVID-19 infection and/or are not suitable for community remote assessment. Several COVID-19 risk scores have been developed. QCOVID provides a risk of mortality considering patients’ existing risk factors but does not include acute signs and symptoms. ISARIC 4C Deterioration model has been specifically developed for hospital settings. In England, the NHS has implemented the Oximetry @home strategy to monitor patients with acute COVID-19 deemed at risk (older than 64 years old or with comorbidities) by providing pulse oximeters; however, the criteria for monitoring or for escalation of care have not been validated. There is, therefore, the need to develop a risk prediction score to establish COVID-19 patients’ risk of deterioration to be used in the community for both face to face or remote consultation.

*Added value of this study:* We developed and validated two COVID-19 specific risk prediction scores. One to be used in the initial remote assessment of patients with acute COVID-19 to assess need for monitoring (RECAP-GP). The second one to assess the need for further treatment escalation and includes peripheral saturation of oxygen among the model predictors (RECAP-O2). To our knowledge, this is the first COVID-19 specific risk prediction score to assess and monitor COVID-19 patients’ risk of deterioration remotely. This will be a valuable resource to complement the use of oximetry in the community clinical decision-making when assessing a patient with acute COVID-19.

*Implications of all available evidence:* To manage pandemic waves and their demand on healthcare, acute COVID-19 patients require close monitoring in the community and prompt escalation of their treatment. Guidance available so far relies on unvalidated tools and clinician judgement to assess deterioration. COVID-19 specific community-based risk prediction scores such as RECAP may contribute to reducing the uncertainty in the assessment and monitoring of COVID-19 patients, increase safety in clinical practice and improve outcomes by facilitating appropriate treatment escalation.

## Introduction

Since the start of the COVID-19 pandemic in 2020, a priority in health systems globally has been rapidly to develop and validate data-driven algorithms to predict risk and guide care. [1-3] This includes a requirement to accurately distinguish between COVID-19 positive patients who can be safely managed in the community setting and those whose care should be escalated to hospital. In primary care, patients with acute COVID-19 have been typically categorised as ‘reassure and safety net’, ‘monitor’ or ‘admit to hospital’ (referred to as green, amber and red risk categories respectively),[4] based on clinical judgment and consensus on using a few clinical parameters such as heart rate, pulse rate, and peripheral oxygen saturation (SpO2).

Several risk prediction models have been used to support the management of acute COVID-19. However, these were derived and validated in hospital inpatient populations.[5] The QCOVID score, based on primary care data, predicts risk of death from COVID-19 based on demographic factors and pre-existing medical conditions, but it does not predict acute deterioration based on current clinical observations.[6] The NEWS2 score is widely used in UK emergency care as an early warning score for sepsis. However, the parameters in NEWS2 (such as tachycardia, fever and hypotension) are usually very late signs of clinical deterioration; have been found to perform poorly in the acute assessment of suspected COVID-19 in hospital inpatients,[7] and have not been evaluated outside hospital.[8] As initial patient contacts with health services are increasingly carried out remotely, a score was needed that could be administered over the telephone or other remote means.[9] Instead of vital signs, such a score could be based on clinical symptoms which the patient or a relative could assess (e.g., perceived breathlessness or confusion). A Delphi study (with qualitative and survey components) using 112 primary care clinicians and 50 patients derived a set of data items comprising symptoms and vital signs that might be included in a putative ‘RECAP: Remote Covid Assessment in Primary care’ prediction model. [10] Templates for collection of these RECAP data elements (known as RECAP-V0), using appropriate SNOMED clinical terms, were developed for Electronic Health Record (EHR) systems so that a model could be derived and then validated.

This study aimed to develop and validate two prediction models: firstly, a score incorporating observable vital signs (heart rate, temperature, respiratory rate, and oxygen saturation); secondly, a score for use when these parameters cannot be measured for lack of equipment or patient familiarity. Two cut-off values, for need of monitoring (green/amber) and consideration of hospital admission (amber/red), for acute COVID-19 were derived and validated.

## Methods

### Study design

RECAP, a prospective cohort and observational study, recruited patients presenting in primary care with symptoms of acute COVID-19. Patients were followed for 28 days from onset of symptoms to determine the occurrence of COVID-related hospital admissions. The previously published study protocol and statistical analysis plan are summarised below. [11, 12]

### Data sources and settings

To allow for parallel derivation and validation of the RECAP scores on different cohorts, we used four different UK primary care settings as shown in Figure 1 below.

**Figure 1.**
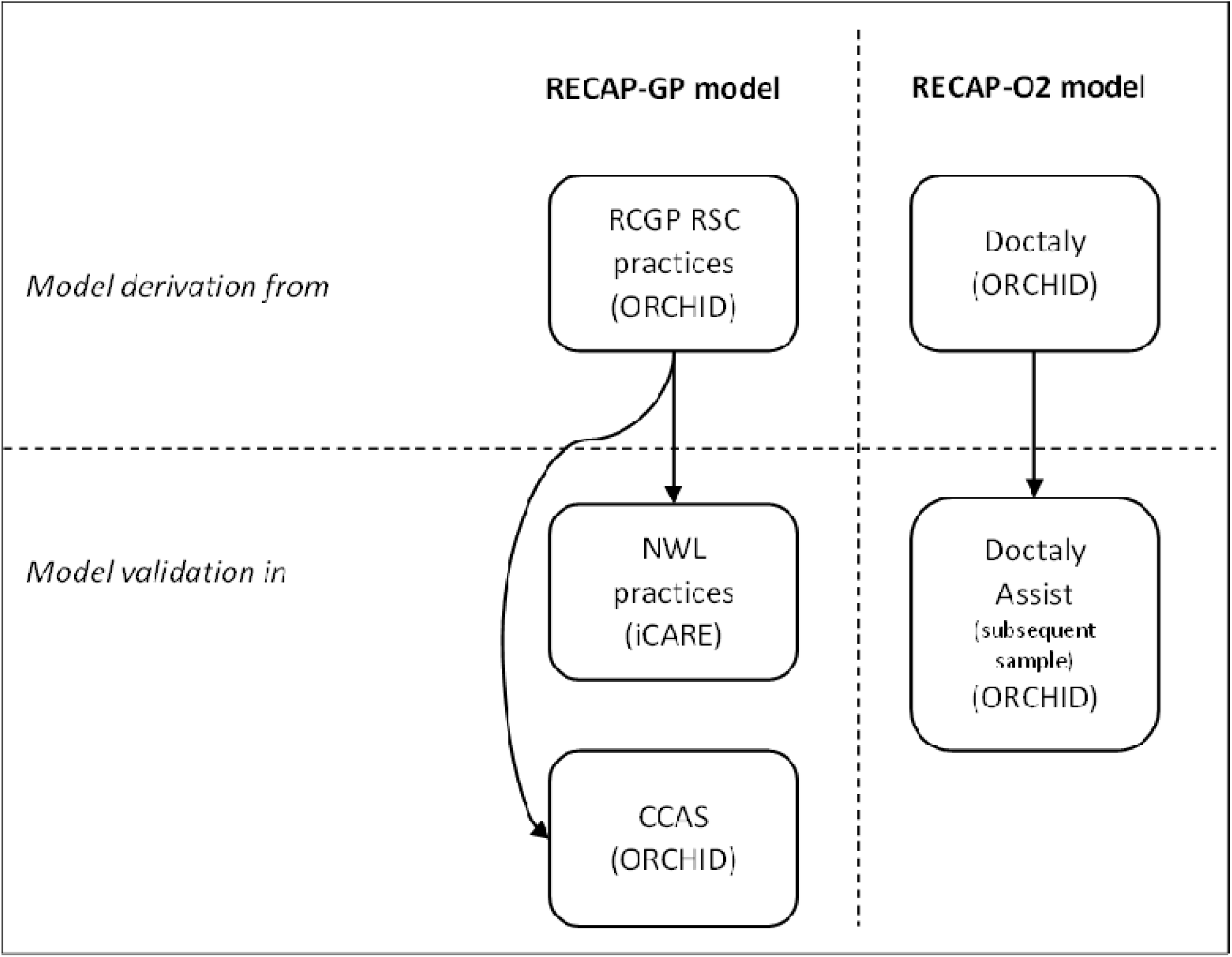
Settings used for derivation and validation of the RECAP scores. RCGP RSC: Royal College of General Practitioners Research and Surveillance Centre. NWL: North West London. ORCHID: Oxford-Royal College of General Practitioners Clinical Informatics Digital Hub environment. iCARE: Imperial Clinical Analytics, Research and Evaluation environment, CCAS: NHS111 Covid Clinical Assessment Service.

### Data

We used primary data on patients’ signs and symptoms collected in the community at the point of consultation linked to secondary data on hospital outcomes. For data collection the RECAP-V0 electronic template, with selected SNOMED codes [10] was used in EHR systems (EMIS, TPP SystmOne and Adastra) and completed by clinicians when assessing patients with signs and symptoms of COVID-19. Additional data on comorbidities, ethnicity, age, and sex, that were included in the development of the risk prediction model were available from the EHR. The Doctaly-Assist platform collected RECAP-V0 data elements via patient self-report (see below). Since the clinical question that needs to be supported is ‘does this patient require care escalation?’, hospital admission i.e, escalation of care from the community, defined as a night’s hospital stay, was the main outcome to be predicted by the model. To derive outcomes, all data were linked with Hospital Episode Statistics (HES) on admissions from NHS Digital in a Trusted Research Environment (TRE), Oxford-Royal College of General Practitioners Clinical Informatics Digital Hub (ORCHID) at Oxford University, or in iCARE (Imperial Clinical Analytics, Research and Evaluation) environments at Imperial College NHS Trust, where data was linked to the NWL ‘SitRep’ data on COVID-19 admissions. A study-specific SNOMED code carrying the RECAP National Institute of Health Research Portfolio Number was used to identify the relevant records for the study in iCARE and ORCHID.

### Data collection in general practice (NWL and RSC)

NWL and Royal College of General Practitioners (RCGP) Research and Surveillance Centre (RSC) [13, 14] primary care practices completed the RECAP-V0 electronic template in EMIS or SystmOne and captured the verbal consent of patients upon completion of the template. Data from NWL practices are routinely included in the ‘Whole Systems Integrated Care’ record and data were accessed in the iCARE system. Data for the RSC practices are routinely extracted and made available for analysis in ORCHID.

### Data collection by COVID-19 Clinical Assessment Service (CCAS)

The CCAS was commissioned from South Central Ambulance Service by NHS England and Improvement to conduct COVID-19 patient remote assessment. A version of the RECAP-V0 electronic template was deployed for use by CCAS clinicians in the Adastra system. Consent was managed in the same way as in primary care practices. Data was transferred to ORCHID for linkage and analysis.

### Data collection by patients using Doctaly Assist

The Doctaly Assist platform is a service provided to patients in South East (SE) London to monitor patients with COVID-19 through a smartphone and with provision of pulse oximeter. [15] Patients are onboarded to a platform which interacts through WhatsApp (see workflow in supplementary material) to ask patients questions based on RECAP-V0 items. [15-17] Data collected were also transferred to ORCHID. Patient consent was not required in this case since data access was granted under Health Service (Control of Patient Information) Regulations 2002 (COPI) notice.

### Sample size calculation

We estimated a minimum sample size of 1,317 participants for model development and 1,400 for model external validation assuming 10% hospitalisation rate for COVID-19, a maximum of 24 predictor variables, a binary outcome (hospital admission), and a minimum 85% model specificity on validation. We aimed to recruit at least 2,880 participants per setting assuming 5-6% loss to follow-up. [12] Patients were recruited according to the following inclusion criteria: having symptoms of acute COVID-19 (within 14 days of onset of symptoms) based on clinical judgment, being 18 years of age or older, and able to provide informed consent (except for Doctaly Assist).

### Statistical analysis

All real-world studies are prone to data missingness, indeed the degree of missingness is a test of suitability of an item for use in clinical settings. The extent of missing data for each variable (outcome and predictors) was assessed on degree of missingness, patterns (at random or not at random), and possible reasons. If the degree of missingness was above 50%, the predictor variable was excluded from the model. If the degree of missingness was less than 50%, the data were imputed using multiple imputation chain equations (MICE). The outcome variable (i.e., hospital admission) was used for the imputation of the predictor variables. We used regression for continuous variables (normally distributed or transformed), and logit or ordinal logit for categorical variables. A total of five imputations were performed and aggregated based on Rubin’s rules.[18] We compared observed and imputed values, especially for variables for which the fraction of missing data was large. A summary of the full methods is provided in the Supplementary Materials and published statistical analysis plan [12]).

A probabilistic risk prediction based on a multivariable logistic regression model, including the variables in RECAP-V0 as factors, was performed for the RSC dataset (model termed RECAP-GP), and separately for the Doctaly Assist dataset (model termed RECAP-O2). The models allow estimation of the likelihood of a particular patient with a COVID-19 diagnosis being admitted to hospital with COVID-19 within 28 days of symptom onset. Variables were checked for independence from each other by including in the model interaction terms between age and respiratory rate where available. When identifying COVID-19 as a cause of admission, we searched for COVID-19 ICD-10 codes, U071 or U072, as first, second or third cause of admission in HES data. COVID-19 as second or third cause were included if the first cause of admission was pneumonia, dyspnea, pulmonary embolism, or chest pain. [19] Multivariable regression models were constructed including all the parameters from the RECAP-V0 template, and their performance assessed to check for significance. Elements of the RECAP-V0 that were constructed using alternative codes with levels of severity (See Table 2) had that relationship maintained in the model. Using backward elimination, a model using only the predictor factors that have shown to be statistically significant with a P value <0.05 was obtained. Internal model validation was conducted using bootstrapping and receiver operating characteristic (ROC) curves plotted. The model performance in sub-populations by age (above and below 65) and by sex was investigated by comparing the diagnostic measures Akaike Information Criterion (AIC), C-Statistic, and visual inspection of the ROC curves. Selection of upper and lower cut-points for creating a Red-Amber-Green categorisation of patients was based on clinical considerations of risk and model performance and consensus or the research team. External validation of the RECAP-GP model was conducted using data separately from NWL primary care practices and CCAS to verify the specificity of the model predictions, as well as the sensitivity, negative predictive value (NPV) and positive predictive value (PPV). The RECAP-O2 model was validated using a subsequent cohort of subjects monitored by the platform. [12]

## Results

The recruitment of patients by practices in NWL and RCGP RSC ran from 1 October 2020 to 28 February 2021. A total of 4,278 patients (NWL=2,415; RCGP RSC=1,863) were recruited by 170 practices (NWL=103; RSC=103). CCAS was actively recruiting from 15 March 2021 to 23 May 2021 and enrolled 2,674 patients. SE London’s Doctaly Assist has provided records for a total of 4,045 patients (1,948 for model development collected between 26 November 2020 and 5 May 2021 (Doctaly-1 dataset); 2,085 for model validation between 8 May 2021 and 26 October 2021 (Doctaly-2 dataset)). The patient populations are described in Table 1. While mean age and sex were similar across the four cohorts, there was a higher proportion of non-white ethnicity in London cohorts, i.e, NWL and Doctaly samples.

**Table 1.** Summary characteristics of demographic data with degree of data completeness.

|  | RCGP RSC<br>(N=1863) |  | NWL<br>(N=2415) |  | CCAS<br>(N=2674) |  | Doctaly-1<br>(N=1948) |  | Doctaly-2<br>(N=2085) |  |
| --- | --- | --- | --- | --- | --- | --- | --- | --- | --- | --- |
|  |  | Complete<br>(%) |  | Complete<br>(%) |  | Complete<br>(%) |  | Complete<br>(%) |  | Complete<br>(%) |
| Mean age<br>(Years)<br>(S.D) | 49<br>(17.7) | 100% | 46<br>(18.3) | 100% | 42<br>(15.9) | 100% | 44<br>(13.1) | 100% | 39<br>(11.9) | 100% |
| Sex (%<br>Female) | 57% | 100% | 56% | 100% | 55% | 100% | 64% | 100% | 61% | 100% |
| Ethnicity<br>(% non-white) | 33.5<br>% | 93.5% | 74% | 91.4% | 78.7% | 85.6% | 68.5% | 92.5% | 51.1<br>% | 96.8% |

**Table 2.** Predictor variables considered for inclusion in the model and whether they were included following missingness assessment. NA - not available * These items contain a proposed severity scale in RECAP-v0 and were captured in the EHR templates as a drop-down list of alternatives with appropriate per item SNOMED coding. This ordering was carried through to the analysis. ** Extracted from GPs EHR systems. *** Collected from the caller as part of the NHS111 pathway.

|  | RCGP RSC<br>(RECAP-GP Model) |  | NWL<br>(RECAP-GP external validation) |  | CCAS<br>(RECAP-GP external validation) |  | Doctaly-1<br>(RECAP-O2 model) |  | Doctaly-2<br>(RECAP-O2 external validation) |  |
| --- | --- | --- | --- | --- | --- | --- | --- | --- | --- | --- |
| Parameter | % Completeness | Considered in model | % Completeness | Considered in model | % Completeness | Considered in model | % Completeness | Considered in model | % Completeness | Considered in model |
| Shortness of breath * | 86% | Yes | 87% | Yes | 96% | Yes | 91.7% | Yes | 100% | Yes |
| Feeling feverish/shivers * | 96% | Yes | 78% | Yes | 96% | Yes | 91.5% | Yes | 100% | Yes |
| Temperature (Observed) | 24% | No | 7% | No | 27% | No | 43% | No | 45% | No |
| Fatigue * | 92% | Yes | 87% | Yes | 91% | Yes | 91.4% | Yes | 100% | Yes |
| Acute cognitive decline * | 82% | Yes | 70% | Yes | 0% | No | 91.4% | Yes | 100% | Yes |
| Time from first symptoms (days) | 93% | No | 32% | No | 96% | No | 99% | Yes | 100% | Yes |
| Respiratory rate | 15% | No | 7% | No | 22% | No | 72% | Yes | 89% | Yes |
| Heart rate | 22% | No | 21% | No | 42% | No | 70% | No | 1% | No |
| Oxygen saturation at rest | 15% | No | 16% | No | 13% | No | 70% | Yes | 62% | Yes |
| Oxygen saturation after 40 steps | 6% | No | 8% | No | 3% | No | 66% | Yes | 57% | Yes |
| Muscle aches | 48% | No | 39% | No | 52% | No | NA | No | NA | No |
| Trajectory of breathlessness * | 45% | No | 52.5% | No | 79% | No | 1% | yes | 0% | Yes |
| Diabetes mellitus | 100% ** | Yes | 100% ** | Yes | 100% *** | Yes | NA | No | NA | No |
| Hypertension | 100% ** | Yes | 100% ** | Yes | 100% *** | Yes | NA | No | NA | No |
| Coronary health disease | 100% ** | Yes | 100% ** | Yes | 100% *** | Yes | NA | No | NA | No |
| Chronic Kidney disease | 100% ** | Yes | 100% ** | Yes | 100% *** | Yes | NA | No | NA | No |
| Age | 100% ** | Yes | 100% ** | Yes | 100% |  | 100% |  | 100% |  |
| Sex | 100% ** | Yes | 100%** | Yes | 100% |  | 100% |  | 100% |  |
| Ethnicity | 93.5% ** | Yes | 91.4%<br>** | Yes | 83.6% |  | 92.5% |  | 3.2% |  |

The RCGP RSC data and the Doctaly-1 datasets were used for model development (Figure 1). The models were subsequently validated in the NWL GP and Doctaly-2 data respectively, as described in the statistical analysis plan (SAP). [12] Table 2 outlines all the model predictor variables considered, and whether they were included in the model following assessment of patterns of missing data.

COVID-related hospitalisation rates were similar in all the datasets except Doctaly-2, 4.4% in the RSC 3.8% in NWL 3.1 % in CCAS, 3.3% in Doctaly-1, but only 0.9% in Doctaly-2 cohort, probably probably because of higher vaccination rates in the Doctaly-2 cohort.

All continuous data were found to be sufficiently normally distributed by visual inspection and the pattern of missingness for each variable was random. MICE was performed on 15 iterations to allow for convergence of the imputation chains. The frequency distributions of the variables in the imputed dataset were compared to the original dataset variables visually and found to be similar and not requiring further transformation (full details in Supplementary Materials).

### Model 1 (RECAP-GP): RCGP and NWL primary care practice data

The RECAP-GP model was built using the RCGP RSC data. The predictor variables used in the final model were sex, age, history of hypertension, degree of breathlessness, and temperature symptoms. (Coefficients and p values shown in Table 3). Fatigue, confusion, ethnicity, body mass index (BMI), diabetes, coronary heart disease, and chronic kidney disease were excluded after backward elimination (p>0.05). The model showed good performance to distinguish between risk levels (AUC=0.802) (Figure 2). There was no significant difference in the performance of the model when stratified for sex and age, although the prevalence of patients in the >65 years group was lower (19%) than those under 65, it was in line with the English population (18.5%)

**Table 3:**
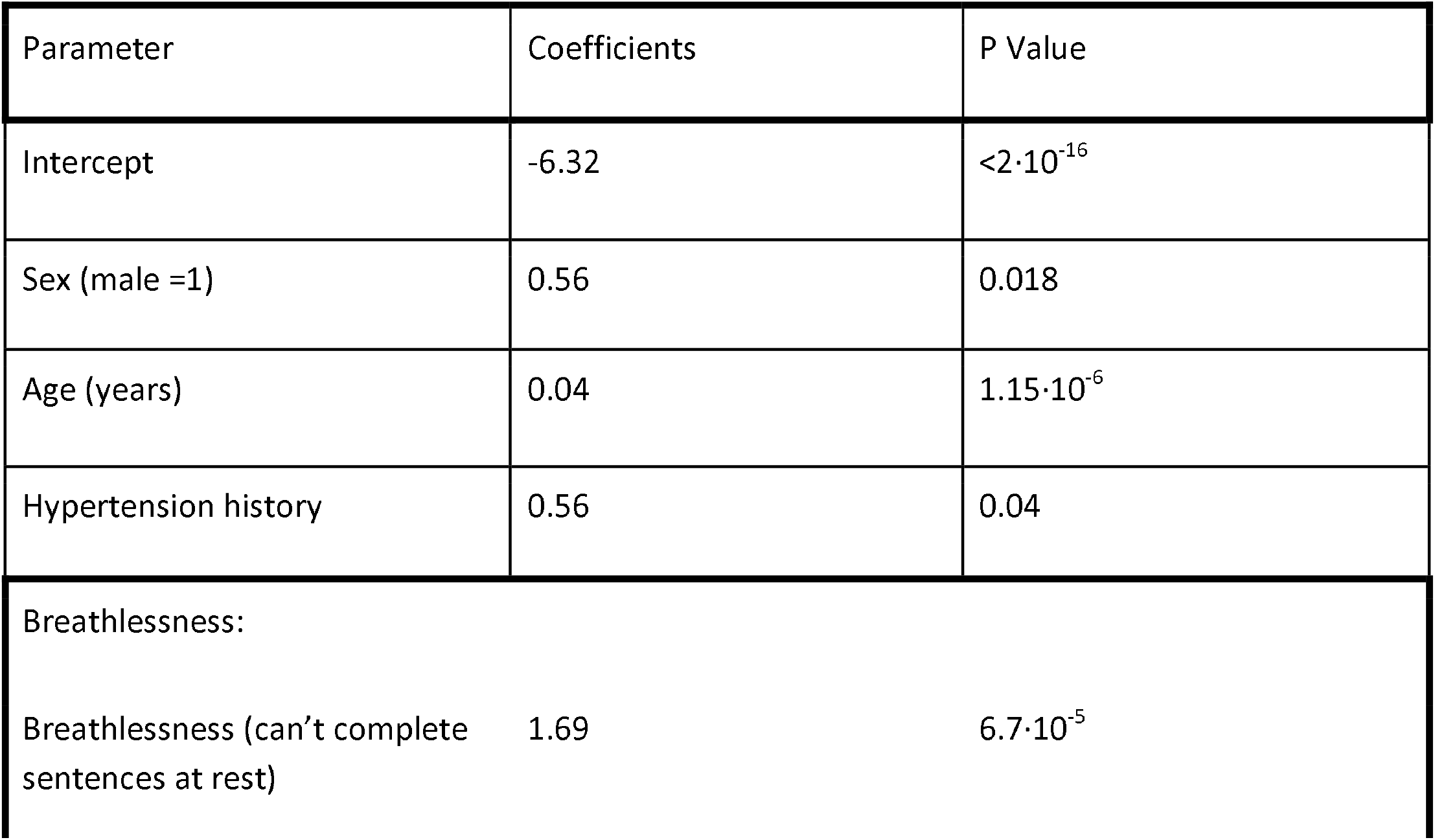
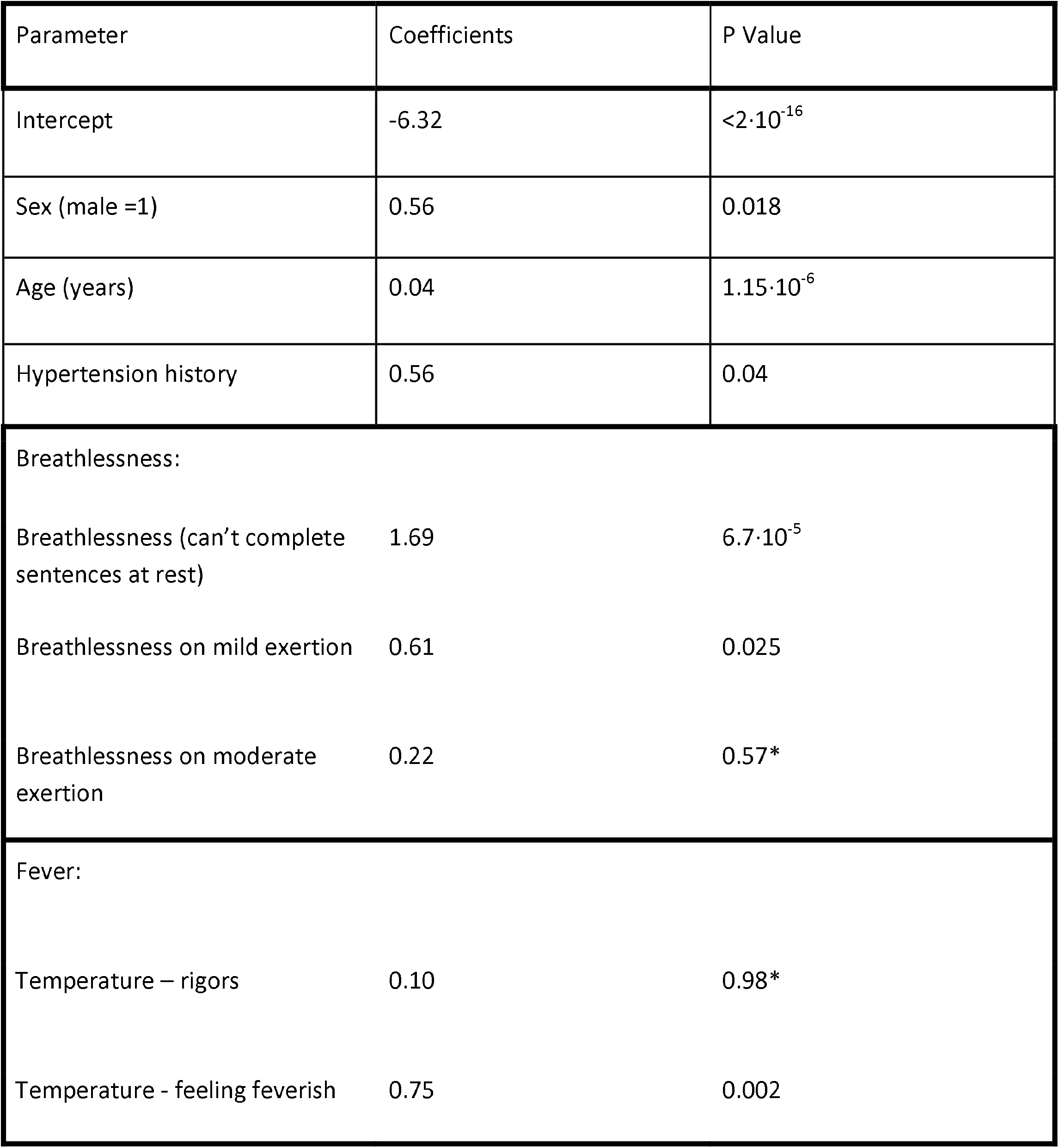
The RECAP-GP model. *For fever and breathlessness severity all levels were included if one level was significant. Absence of hypertension, breathlessness and fever are the base coefficients in the logistic regression, set to zero and not shown.

**Figure 2.**
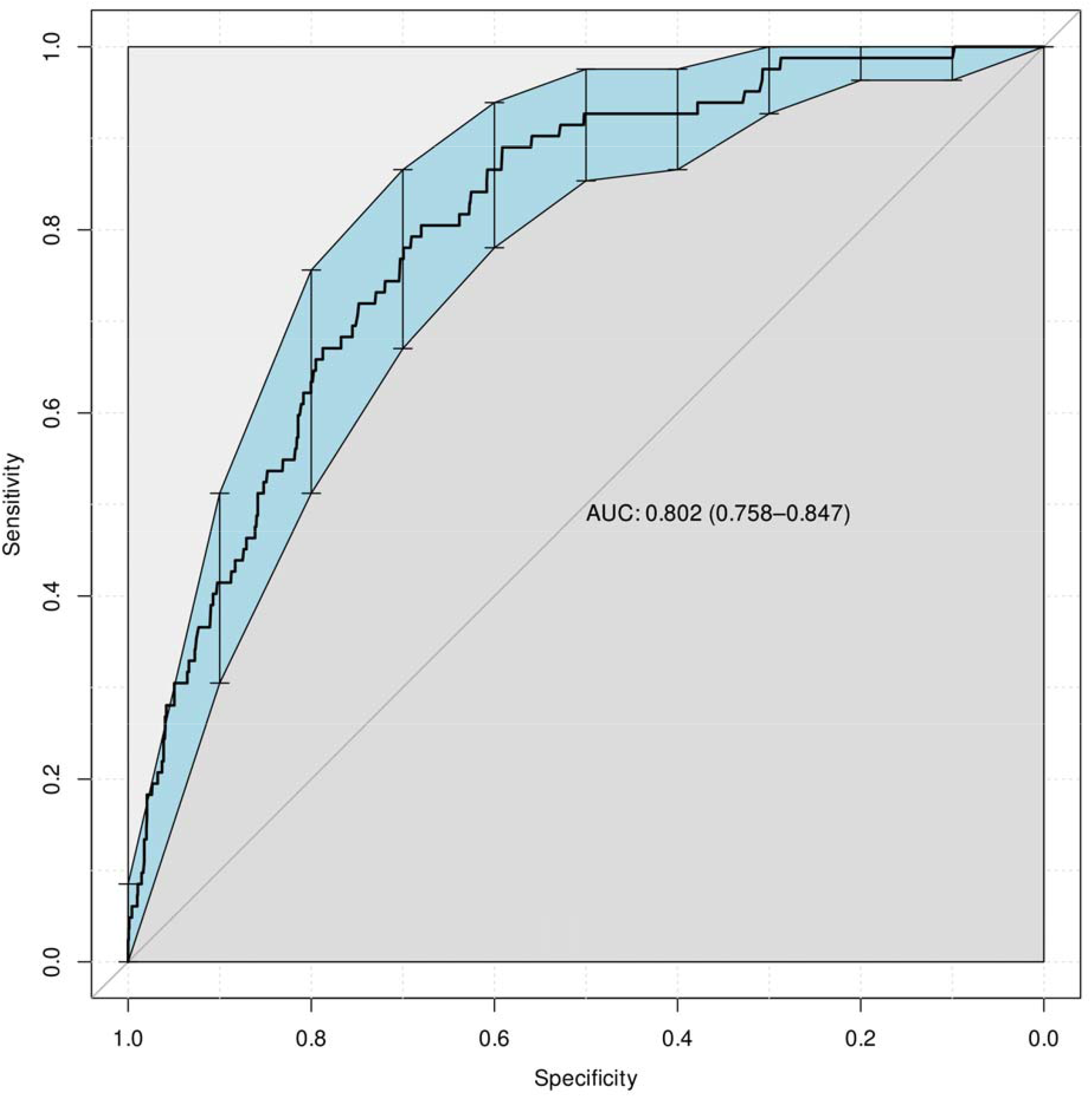
ROC curve of the RECAP-GP model performance following bootstrapping for internal validation along with model diagnostic measures obtained as part of model calibration and performance assessment. (AUC: area under the curve, AIC: Akaike Information Criterion)

The cut-off points for the green, amber, and red risk groups were chosen by the research team clinicians (BD, SdeL, ErM, EmM, ElM, TG, AE-G), before validation, using the specificities and sensitivities obtained from the ROC curve shown in Figure 2. The selected cut-off points and related specificity, sensitivity and interval likelihood ratios (ILR) are shown in Table 4. We opted for maximising model sensitivity (90%) for the low to moderate risk threshold to ensure all patients needing monitoring were in the amber group and maximising specificity (90%) in the moderate to high risk threshold to try to limit the number of unnecessary hospital admissions from the amber group.

**Table 4.**
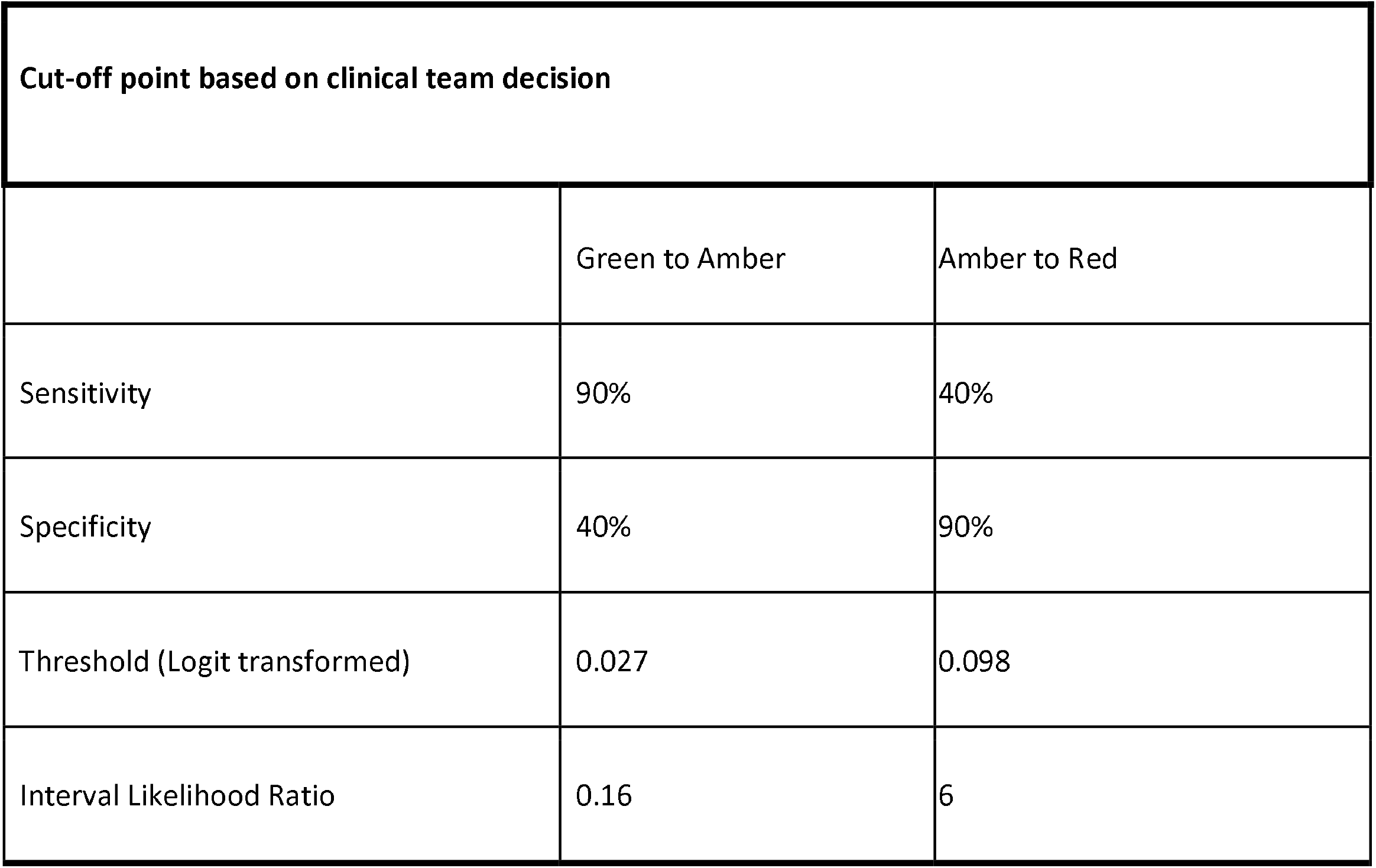
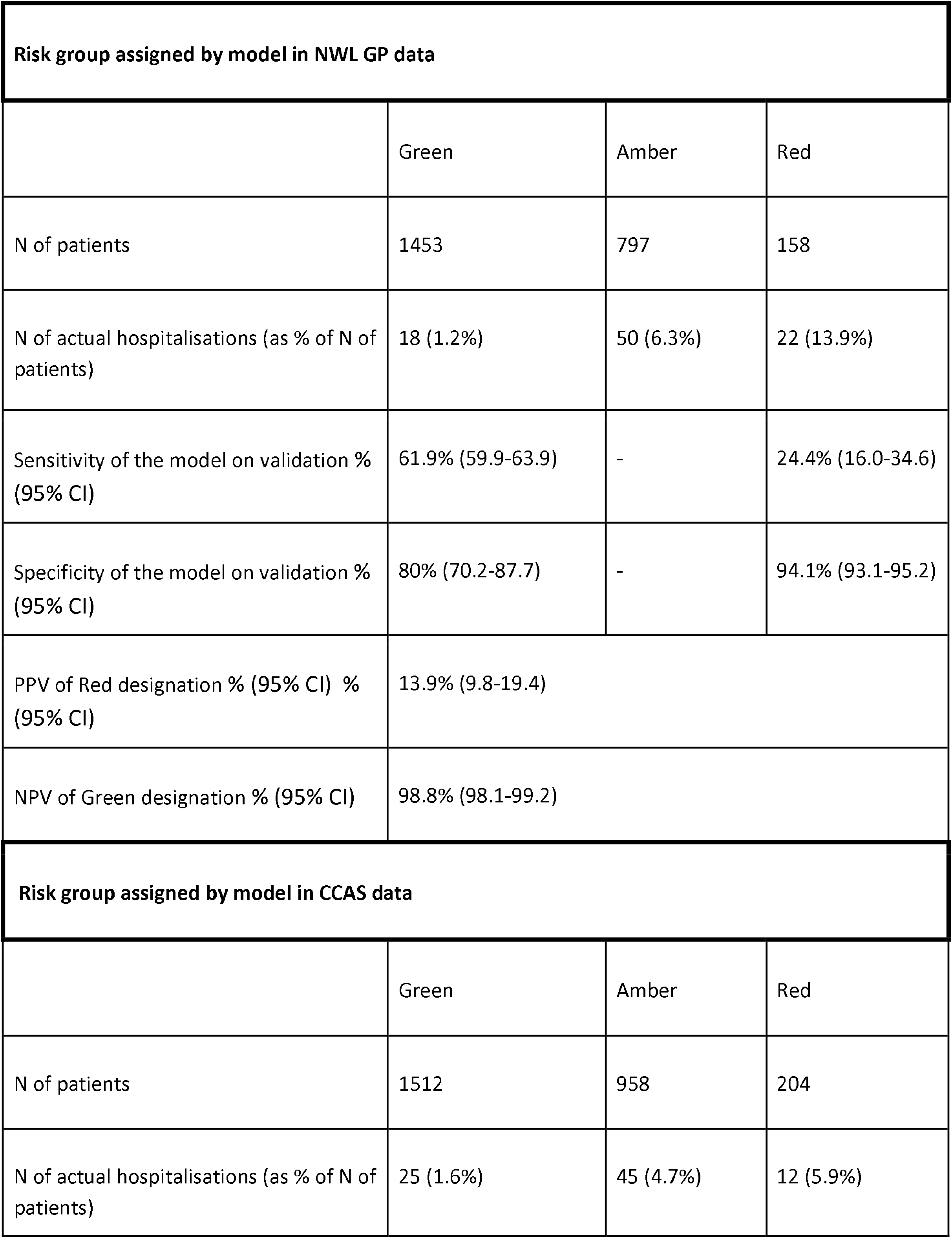
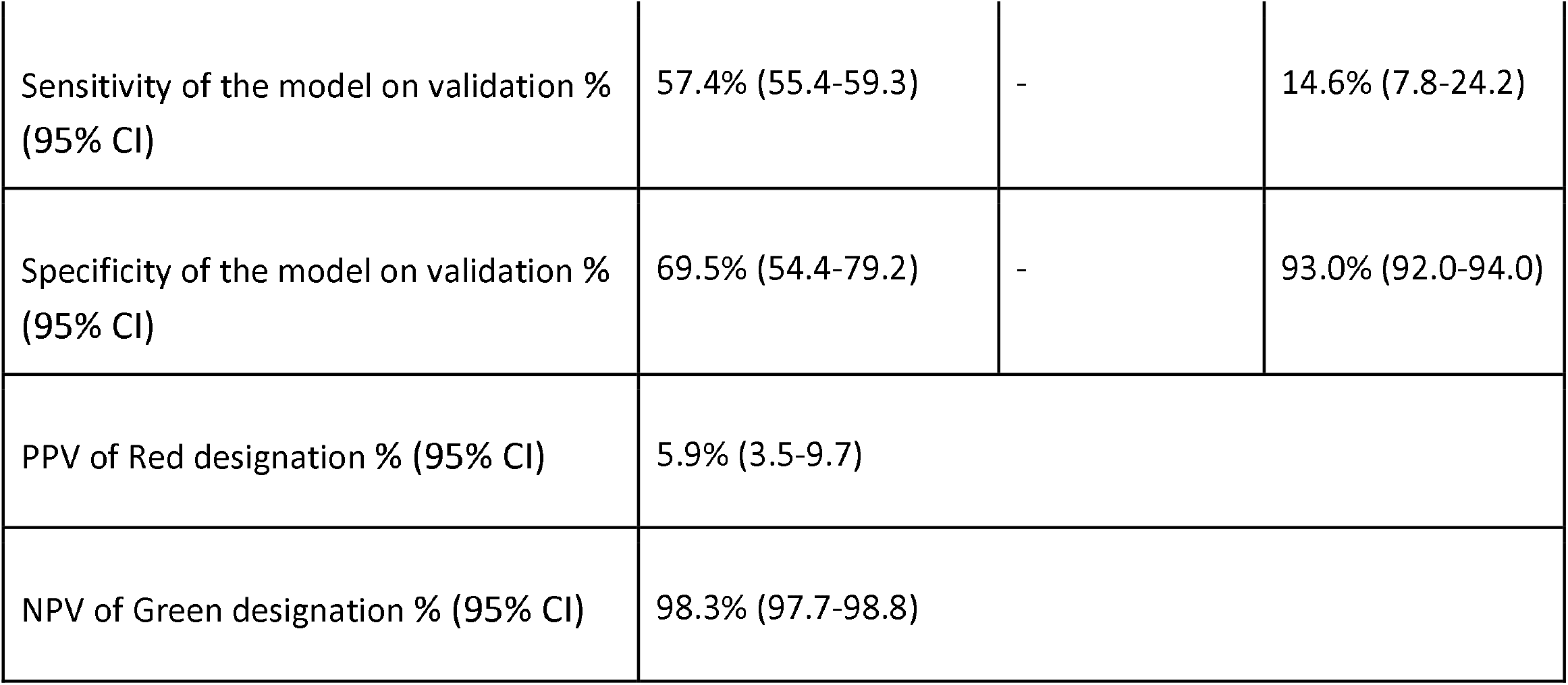
Cut-off points for the RECAP-GP model along with sensitivity, specificity, negative and positive predictive values following external validation in the NWL and CCAS data. Positive predictive value (PPV): N of hospitalisations in red group /N of patients in red group. Negative predictive value (NPP): Number of patients non-admitted in green group/N of patients in green group

For external validation, the prediction model was run using both the NWL data and the CCAS data separately. We decided not to build a separate CCAS model as originally intended, as the data was similar to the GP practice data, and lower than initially expected (2020) admission rates would have limited power. The selected cut-off points were used to assign risk categories to patients as shown in Table 4, along with the observed model sensitivity, specificity, NPV and PPV. As true negative and true positive patients for the amber group cannot be defined, only the number of final hospitalisations is noted in the results. In the NWL GP data, the probability of being categorised as a low risk (green) patient and not needing admission (i.e., NPV) was high (98.7%), but the probability of being in the high risk (red) group and being admitted (i.e., PPV), was low (13.9%). In the CCAS data, the NPV was 98.3%, equivalent to the GP data, but the PPV was lower (5.9%), the confidence intervals just separated from the NWL validation.

### Model 2 (RECAP-O2): Doctaly-1 and Doctaly-2

Given the predicted availability of larger numbers of Oxygen Saturation (SpO2) readings in the Doctaly Assist data, we planned a second model using the Doctaly-1 dataset. Predictor variables used in the final model were age, degree of breathlessness, fatigue, and SpO2 at rest (coefficients and p-values shown in Table 5). Sex, ethnicity, temperature, acute cognitive decline, days since onset of symptoms, respiratory rate, and trajectory of breathlessness were excluded after backwards elimination (significance >0.05%). SpO2 after activity was found to be co-linear with SpO2 at rest in the model and was thus excluded from the final model.

**Table 5:** The RECAP-O2 model. Note ‘scales’ of severity are included if one element is significant and only the most severe level is used in the model. *For fatigue and breathlessness severity all levels were included if one level was significant. Absence of hypertension, breathlessness and fatigue are the base coefficients in the logistic regression, set to zero and not shown.

| Parameter | coefficient | P Value |
| --- | --- | --- |
| Intercept | 25.0 | $3.85 \cdot 10^{-7}$ |
| Age (years) | 0.04 | 0.0002 |
| Breathlessness: |  |  |
| Breathlessness – Feeling Uncomfortable to breathe | 0.92 | 0.03 |
| Breathlessness when walking* round the room | -0.43 | 0.26 |
| Fatigue: |  |  |
| Fatigue - Difficult to wake up | 1.5 | 0.068 |
| Fatigue - Too tired to do usual activities | 1.23 | 0.0007 |
| Oxygen saturation at rest % (0-100%) | -0.33 | $9.8 \cdot 10^{-11}$ |

The model was internally validated and calibrated through bootstrapping. Figure 3 shows the ROC curve along with the model diagnostic analysis. The AUC was 0.843 and AIC 354, which suggests good model performance. Notably the bottom left of the ROC curve has a steeper slope than the RECAP-GP model, indicating better discrimination in sicker patients. The model performance in sub-populations by age (>65) and sex was investigated and was not found to be significantly different.

**Figure 3.**
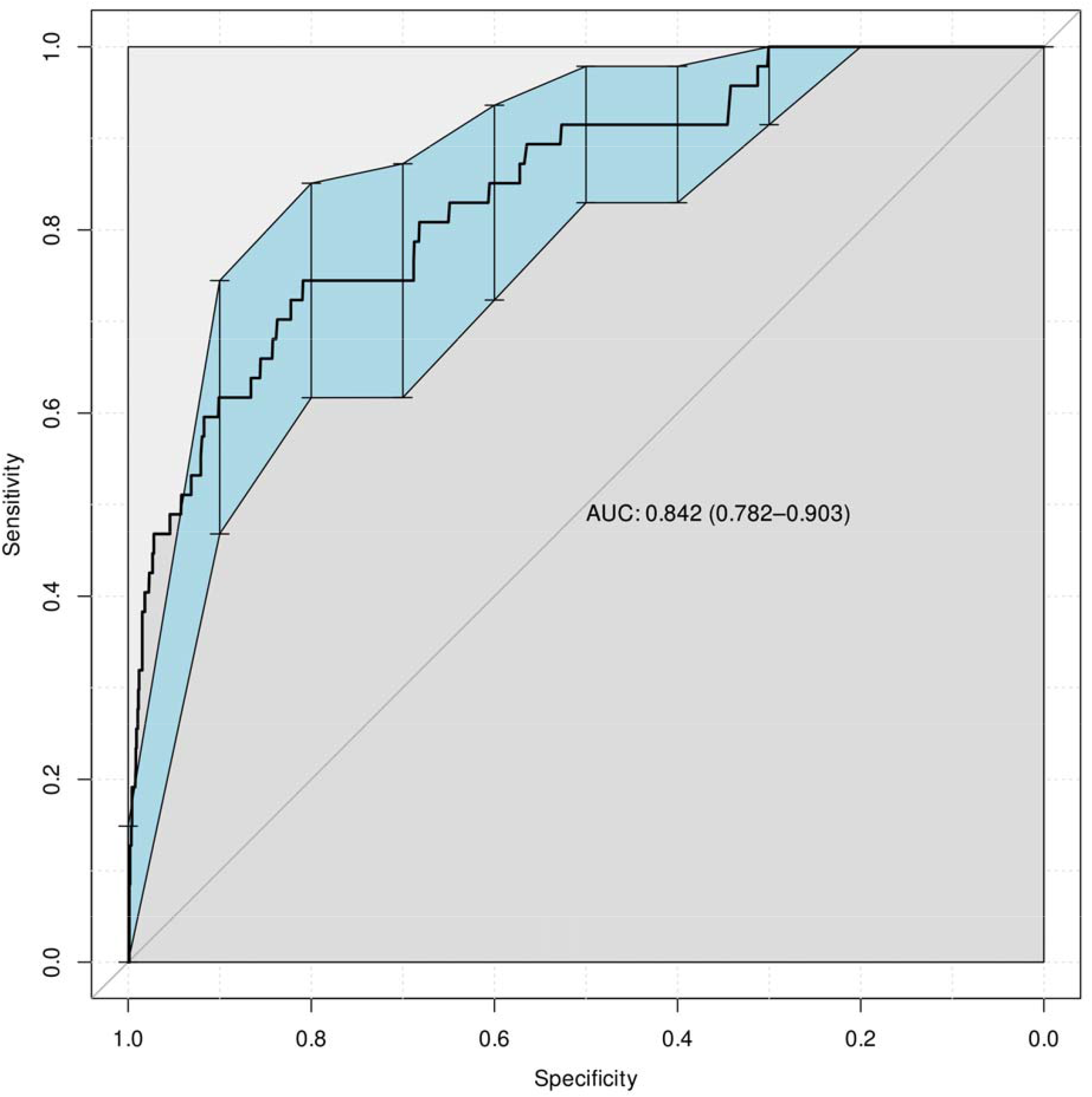
ROC curve of the RECAP-O2 model performance following bootstrapping for internal validation along with model diagnostic measures obtained as part of model calibration and performance assessment.

For external validation the RECAP-O2 model was run using the Doctaly-2 data. Likewise, as seen in Table 6, the cut-off points were chosen by the clinical team based on the ROC curve in Figure 3 and were used to assign risk categories to patients. Table 6 shows the selected cut-off points, related specificity and sensitivity, and the actual sensitivity and specificity, negative and positive predictive values of the RECAP-O2 model following external validation. Although NPV of green designation is slightly higher than RECAP-GP (99.4%), PPV of red designation is lower (8.8%), on account of the lower rates of admission in the Doctaly 2 data (the population now having the majority vaccinated), which means lower probability of being admitted when categorised as a high risk patient.

**Table 6:**
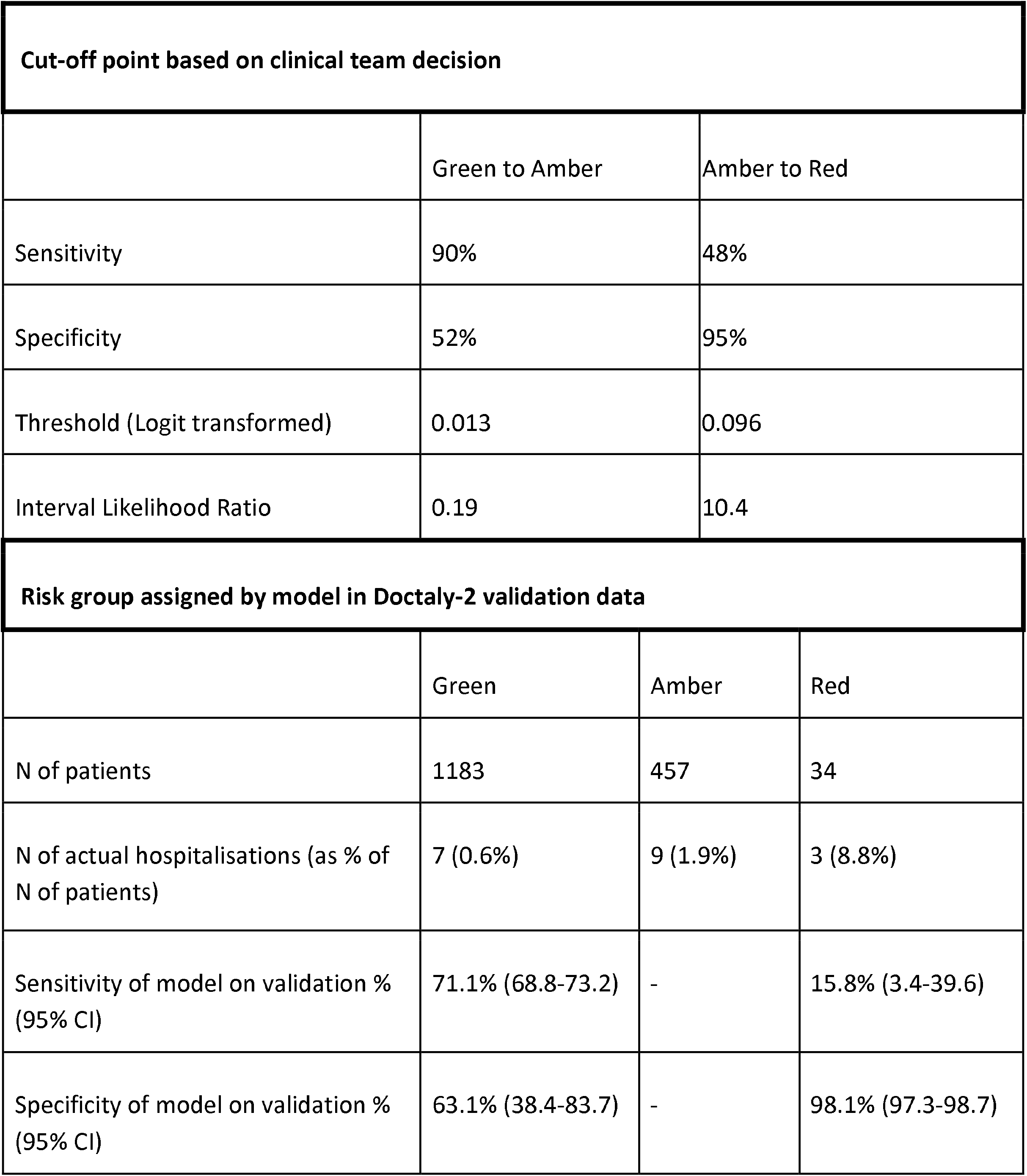
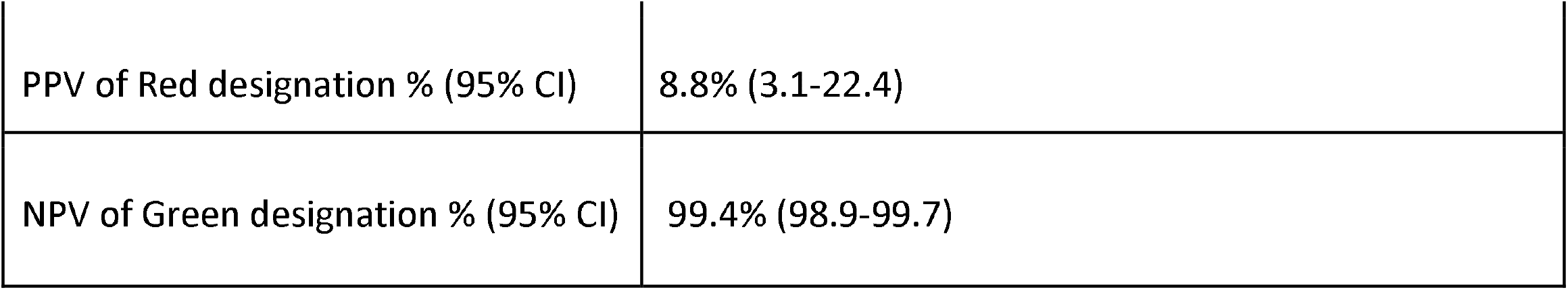
Cut-off points for the RECAP-O2 model (values chosen by the clinical team) along with sensitivity, specificity, negative and positive predictive values following external validation. Positive predictive value (PPV:) N of hospitalisations in red group /N of patients in red group Negative predictive value (NPP): Number of patients non-admitted in green group/N of patients in green group

| Cut-off point based on clinical team decision |  |  |  |
| --- | --- | --- | --- |
|  | Green to Amber | Amber to Red |  |
| Sensitivity | 90% | 48% |  |
| Specificity | 52% | 95% |  |
| Threshold (Logit transformed) | 0.013 | 0.096 |  |
| Interval Likelihood Ratio | 0.19 | 10.4 |  |
| Risk group assigned by model in Doctaly-2 validation data |  |  |  |
|  | Green | Amber | Red |
| N of patients | 1183 | 457 | 34 |
| N of actual hospitalisations (as % of N of patients) | 7 (0.6%) | 9 (1.9%) | 3 (8.8%) |
| Sensitivity of model on validation % (95% CI) | 71.1% (68.8-73.2) | - | 15.8% (3.4-39.6) |
| Specificity of model on validation % (95% CI) | 63.1% (38.4-83.7) | - | 98.1% (97.3-98.7) |

*Table 6: Cut-off points for the RECAP-O2 model (values chosen by the clinical team) along with sensitivity, specificity, negative and positive predictive values following external validation.
|  |  |
| --- | --- |
| PPV of Red designation % (95% CI) | 8.8% (3.1-22.4) |
| NPV of Green designation % (95% CI) | 99.4% (98.9-99.7) |

## Discussion

Assessment of severity of COVID-19 in the community is crucial to pandemic management worldwide. Our study provides a derivation and real-world validation of two risk scores specifically designed for COVID-19 patients in the community. The RECAP-GP model includes degree of breathlessness, temperature symptoms, presence of hypertension, sex, and age as hospital admission predictors, and can be used when a pulse oximeter is not available to the patient. This model provides a good prediction for risk of non-admission in the lowest risk group, e.g., from 2,415 patients in NW London, 60.1% were assigned to the green group with a NPV of 98.7%. The model performs less well at differentiating amber from red groups, with only 22 out of 158 red patients needing admission (PPV 13.9%). When validated in the CCAS data the RECAP-GP model performs slightly less well on PPV. However, performance on the green/amber cut is within the NWL confidence intervals with 57% patients being safely reassured. The RECAP-O2 model included degree of breathlessness, fatigue, SpO2 at rest, and age as predictors. This model can be used if pulse oximeters are available, such as in moderate risk patients under a monitoring service. Although the improvement of the slope on the ROC curves indicates an Interval LR increasing from 6 to 10 for the amber/red cut point, the model performs less well on validation with a PPV of 8.8%, due to the lower admission rate in the now majority-vaccinated validation set. However, its specificity is good (98.1 %), which supports its use to assess moderate risk patients’ need for admission (only 9 of 457 amber patients required admission).

Much discussion has taken place around the potential role of SpO2 measurement as an early warning for disease severity. In the UK a national strategy for using home pulse oximetry, COVID Oximetry @home, was established during 2020 with the intention of identifying ‘silent hypoxia’ (without breathlessness). UK guidance recommends provision of pulse oximeters, arranged by local health authorities, to monitor patients with symptomatic COVID-19 older than 65 years old or with risk factors. [16] The use of RECAP in the assessment of COVID-19 patients is a valuable addition to this strategy. Our results are aligned with the guidance since age and SpO2 significantly predicts deterioration in our models, but RECAP also considers symptom severity so it can better support clinicians’ judgment on who needs monitoring, particularly for younger patients without comorbidities. Moreover, it is better at identifying need for treatment escalation compared with SpO2 alone, which is only one factor in the final model.

The RECAP models are founded on assessment of need for hospital admission in an observational dataset. In such a design it is impossible to eliminate ‘incorporation’ bias due to the admission decision being partly based on the parameters included in the model (e.g., SpO2). We mitigated this by counting admissions as at least one night in hospital, rather than only a review in the emergency department. Recruitment was conducted contemporaneously via GP practices, CCAS, and the Doctaly Assist service from fall of 2020 to spring of 2021. However, the Doctaly Assist validation dataset was collected during the summer of 2021 as the UK’s COVID-19 vaccination programme covered up to 70% of the adult population. [20] This increasing vaccine coverage may account for the difference in validation of the RECAP-O2 model due to lower admissions being observed. Whilst it is unlikely that SARS-Cov-2 variants and vaccination status will affect the predictive signs of deterioration, these being driven by common pathophysiological processes, the overall lower event rates demand better performance in a model. However, all models should be subject to ongoing surveillance/calibration, especially with rapid changes in variants and vaccines.

In any study relying on data collection during routine practice there will be missing data. It is notable that remote consultations with GPs did not enable collection of vital signs from patients. Health monitoring devices, such as pulse oximeters and thermometers, are rarely available in the community unless patients are being provided with them. Therefore, these observations were largely missing in the GP and CCAS datasets and could not be included in the RECAP-GP model. In contrast, the Doctaly Assist datasets enabled us to assess the predictive value of SpO2, resting and on exercise, heart rate, respiratory rate, and temperature, determining, for the first time, the diagnostic value of Sp02 monitoring in the community.

We took a careful approach to analysis with peer-review and prior publication of the protocol and Statistical Analysis Plan. The use of MICE and bootstrapping for the internal validation are standards in the development of diagnostic models. We also chose several relevant validation datasets. However, lack of availability of EHR data linkage for the Doctaly data limits the ability to include factors such as co-morbidity in the RECAP-O2 model. The age distributions, admission rates, ethnicity, and comorbidities are in line with UK population expectations, except the Doctaly data has less over 75 yr olds than expected. SE London and NW London contain significant populations of black British and South Asian ethnicity, whilst the RSC network is more representative of the UK. [21] This supports the external generalisability of our findings. The thresholds used in the two RECAP scores can be adjusted to better suit local circumstances and, in the future, it may be necessary to consider re-testing a model with vaccination status as a potential predictor. We did not do this as data on vaccination status were not consistently available and policy was changing rapidly during the study.

The RECAP-V0 template developed by the Delphi study contained 10 questions, the validated models contain four and five items only, significantly improving their fitness for use in the clinical setting.[22] The choice of logistic regression modelling means that most EHR systems using SNOMED codes will be able to recreate the RECAP electronic templates and integrate the score calculator into the system. The full models are provided as a downloadable code on Github.

The identification of cut-off points to differentiate between risk levels was based on clinical consensus. We intended to maximise the model sensitivity when deciding on the green-amber risk level (so as not to miss cases requiring monitoring), while intending to identify an amber-red cut-off point specificity that minimises unnecessary hospital admissions. The result is a tool that can be safely used for initial assessment and monitoring if appropriate monitoring mechanisms to identify deterioration are in place for the amber group. We suggest that the RECAP-GP score be used remotely in the initial assessment, without need for patient observations. Following this, if the patient is considered moderate to high risk, they could be provided with a home pulse oximeter for calculation of the RECAP-O2 score to detect deterioration. Given the low PPV of the Red categorisation on the amber/red cut point the RECAP-O2 model will tend to over alert. Care escalation should be considered with reference to national or local pathways, ability to monitor the patient in the community, hospital capacity and shared decision making with the patient.

Much has evolved since the first wave of the COVID-91 pandemic when we conceptualised this study. Mass vaccination has dramatically changed prognosis and oximetry is now much more widely used in the community than it was in early 2020, with ‘at home’ services available in many settings. Yet, new variants are triggering new pandemic waves across the globe, putting services under great strain. We believe that these two scores are likely to be a valuable resource to support clinical judgement, reduce uncertainty and improve safety in triage and monitoring of patients with suspected COVID-19 in health systems worldwide.

## Data Availability

All data produced in the present study are available upon reasonable request to the authors

## Contributors

AEG coordinated recruitment across sites, helped with template development and elaboration of datasets in WSIC/iCARE and ORCHID and drafted the manuscript. DP and FF contributed to the design of the analysis, conducted the analysis and contributed to interpretation of the results. CR supported the ethics submission and amendments, communications with practices during the recruitment period, DPIA, COPI, and CAG applications. ElM and EmM contributed to code and template development, analysis and interpretation of results. BG contributed to the conceptualisation and design of the study, managed the data in iCare, contributed to the analysis and interpretation of the results. ALN contributed to the development of the protocol, helped obtain funding, and contributed to the interpretation of the results. CO managed the data in ORCHID and contributed to the analysis and interpretation of the results. LH contributed to obtaining funding, helped write the protocol and assisted in the management of data collection in practices. RC and JH assisted in the design of the SE London Covid management pathways, helped obtain funding, assisted data collection and interpretation of the analyses. MB, CW and BB coordinated the patient enrollment at CCAS. BD, ErM, S deL and TG developed the protocol and obtained funding. BD supervised the study and acts as corresponding author. All authors contributed to interpretation of results and drafting of the manuscript.

## Data sharing

The models are available for download from the following link: TBA

## Acknowledgements

Doctaly Assist (Phil Tyler and Dr Prad Velayuthan) provided the data extraction and SNOMED coding of their Chat-bot data. Dr Mark Ashworth and Dr Ibi Fakoya of King’s College London evaluated the initial set-up of the Covid monitoring programme in SE London. Eamon O’Doherty of NW London Clinical Commissioning Groups extracted the relevant data tables from WSIC to enable the data analysis in iCare. Sneha Anand at Oxford University managed the governance, data transfer and linkage in ORCHID. Matt Widdows coordinated the development of the RECAP template with Adastra and extracted the relevant CCAS data for analysis. Dr Merlin Dunlop of Ardens assisted in the SNOMED coding of the RECAP-V0 templates and their distribution. Ashnee Dhondee of NWL CRN helped recruit practices to the study.

## Dissemination

The study was presented at the 1º Congresso Brasileiro de Evidências Clínicas na COVID-19 in May 2021 (reference) and at the SAPC ASM and FCI Scientific Conferences both in July 2021. The RECAP-V0 model has informed the development of COVID-19 clinical assessment tools in other countries, such as the online assessment tool set up by the Fiocruz Oswaldo Cruz Foundation in Rio de Janeiro, Brazil (https://redcap.ini.fiocruz.br/surveys/?s=HPHCHDEDHN).

## Ethics

The study was sponsored by Imperial College London and approved by the North West-Greater Manchester East Research Ethics Committee and Health Research Authority in May 2020 (IRAS number: 283024, Research Ethics Committee reference number: 20/NW/0266). The study was badged as an Urgent Public Health Study by the National Institute of Health Research in October 2020.

The WSIC data analysis was undertaken within a research database that was given favorable ethics approval by the West Midlands Solihull Research Ethics Committee (reference 18/WM/0323; IRAS project ID 252449). All data used in this paper were fully anonymized before analysis. iCARE is a Trusted Research Environment and provides access to HRA REC approved anonymised data for research (REF 21/SW/0120, IRAS project ID: 282093)

At Oxford, analysis was undertaken within the secure data processing platform of the Oxford Royal College of General Practitioners Clinical Informatics Digital Hub (ORCHID) trusted research environment (TRE). (DIPA registration number: Z575783X, DARS number: DARS_NIC_431881_N8B0N, https://orchid.phc.ox.ac.uk/index.php/orchid-privacy-notices/).

The access to Doctaly Assist data was granted under COPI notice (and CAG Resolution 5 after expiration of COPI notice in March 2022) and, therefore, patient consent was not required. However, patients could opt-out using the National Opt-Out register.

