## Supplementary material for "Remote Covid Assessment in Primary Care (RECAP) risk prediction tool: derivation and real-world validation studies"

#### Supplementary Materials

#### 1. Methods

##### Selection of outcomes

Hospital Episode Statistics data consists of a set of ICD10 codes for reasons for admission with dates of admission and discharge. The first field is the primary reason for admission as chosen by the clinical coders. To identify COVID-19 related admissions within our cohort, we selected cases that had U07.1, and U07.2 codes (COVID-19 on PCR test or clinical diagnosis respectively), as well as U07.3 (personal history of COVID-19) and U07.4 (current cause of admission likely related to COVID-19) as the primary cause of admission. We also searched for those codes in second, third or further cause of admission and included the cases in our outcomes cohort if the primary cause was respiratory (e.g., viral pneumonia or acute respiratory failure) or cardiac (e.g., pulmonary embolism, chest pains or palpitations) and the admission had taken place within 28 days of acute COVID-19 onset of symptoms. All admission codes were reviewed by the clinical authors. Table 1 depicts the COVID-19 related hospitalisations per cohort.

*Table 1 Hospitalisation per cohort*

| | **RCGP RSC** | **NWL** | **CCAS** | **Doctaly-1** | **Doctaly-2** |
|---|---|---|---|---|---|
| **Total cohort** | 1,863 | 2,415 | 2674 | 1,948 | 1,555 |
| **Total COVID-19 related admissions** | **82** | **92** | **82** | **64** | **18** |
| **First cause** | 79 | 92 | 77 | 52 | 17 |
| **Second cause** | | NA | | | |
| Cardiovascular | 1 | | 3 | 4 | |
| Respiratory | 2 | | 1 | 1 | |
| Other complications | | | 1 | 1 | 1 |
| **Third or other cause** | | NA | | | |
| Cardiovascular | | | | 2 | |
| Respiratory | | | | 2 | |
| Other complications | | | | 2 | |

#### Model development

The assumptions of the model were checked including model specification, goodness of fit, multicollinearity, and the effect of influential observations. Model specification was assessed by looking at the McFadden's $R^2$ value of the model and the AIC (Akaike Information Criterion) to check the overall performance of the model. The presence of outliers and their influence on the fit of the model was assessed by calculating the residual vs leverage of all observations and calculating their Cook's distance. Points identified as outliers were removed from the dataset before model re-calibration. The link function of the models was chosen as the logit function due to the outcome being a binary variable. Individual terms of the final model were assessed for significance by running a chi-squared ANOVA test where each of the terms was added sequentially, and the significance of their contribution assessed relative to the final performance of the model. Continuous variables were assessed for non-linearity by fitting splines through penalized thin plane regression and noting the effective degrees of freedom, as well as changes to model performance compared to fitting a linear model.

The specificity, sensitivity, positive and negative predictive values were calculated.

#### Internal validation

The model was validated internally by bootstrapping to ensure it performed well. The bootstrapping was performed by resampling the original dataset 500 times and pooling the results of the aforementioned diagnostic tests for all models on all bootstrapped datasets. The distributions of the Brier scores and c-index were assessed for normality, and optimism-corrected c-statistic and shrinkage factor was calculated for each model. The goodness of fit of the model was assessed using AUROC and optimism-corrected smoothed calibration curves to determine the performance of the model in predicting outcomes in the bootstrapped cohorts, with the final relationship between the predicted and the observed data being expressed as McFadden's R2 value and the optimism-corrected c-statistic.

To provide better representation of model fit, calibration and discrimination were performed to better evaluate the accuracy of the model. Calibration of the model was achieved by using LOESS-smoothed calibration plots after bootstrapping the data. To analyse discrimination, the data were stratified by age (65 years old as cut off point) and gender to test the model in population subgroups with age and gender for the subgroup definition. Brier scores were used to measure the accuracy of the predictions of the model in the bootstrapped data. A receiver operating characteristic (ROC) curve was plotted to determine the cut off points for the predicted risk that optimises the specificity and sensitivity of the predictor model for red, amber, and green risk groups. The risk groups represent risk of a hospitalisation to assist in managing patients presenting in the community, with green signifying a very low risk, amber being a moderate to high risk, and red being a very high risk. The choice of optimum cut points was a clinical decision based on the shape of the resulting ROC curve. Each clinician in the research team was presented the ROC curve and made decision on the cut-off point independently, followed by a discussion as a team. There are several statistical tools for finding a single cut-off point, but none in use for multiple points. In our view, selecting appropriate points from inspection of the ROC and pre-stated aims of optimal sensitivity or specificity has the highest clinical validity.

#### 2. Analysis

##### RECAP-GP

Figures 1, Figure 3 and Figure 3 below show the age distribution in the RCGP RSC, NWL, and CCAS cohorts respectively that were used to develop and validate RECAP-GP model.

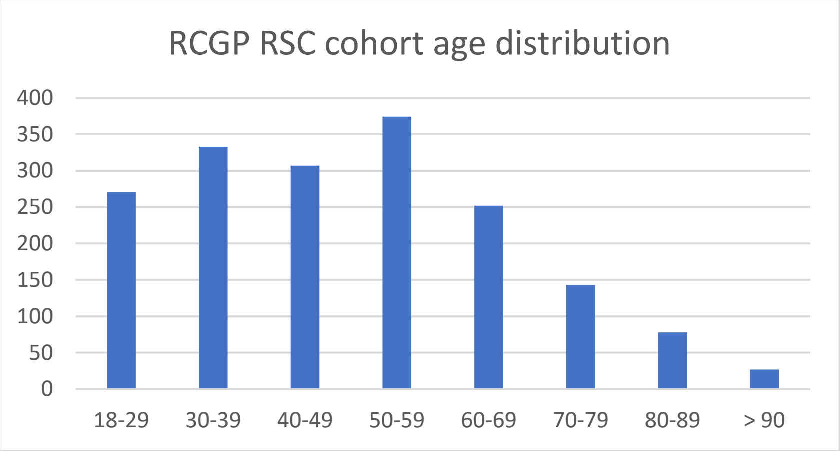


*Figure 1 Age distribution in RCGP RSC cohort*

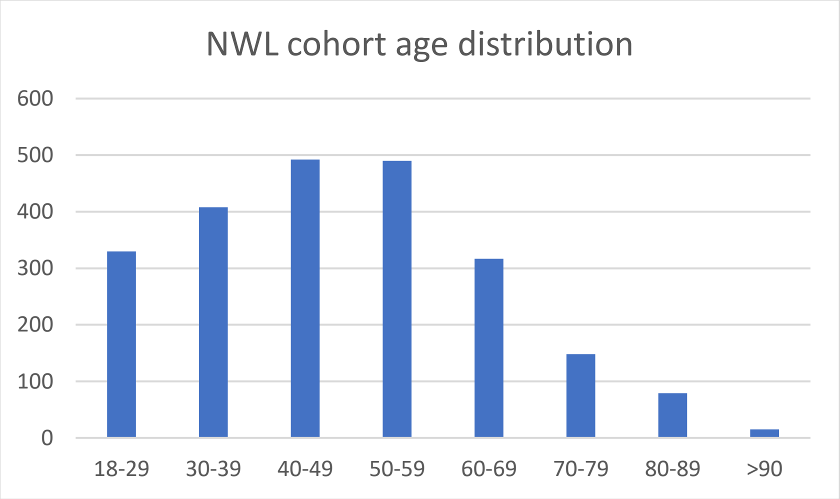


*Figure 2 Age distribution in NWL cohort*

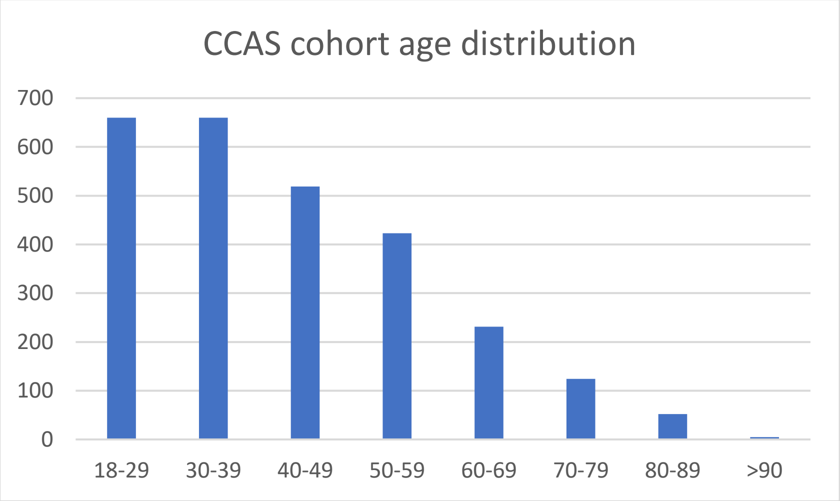


*Figure 3 Age distribution in CCAS cohort*

#### Parameter non-linearity

Potential non-linearity of age and BMI were tested by including them as complex splines fitted through penalised thin plane regression. There was no non-linearity in the fitted splines as dimensionality was optimal at 1 degree of freedom, indicating a linear slope of fit.

#### Parameter interaction

The pre-defined interactions, between age and degree of breathlessness as well as BMI and degree of breathlessness, were investigated assessing their significance and the relative influence on the model given the size of the coefficient fitted in the logistic regression. The p-values indicated (0.8, 0.59, 0.78) that the interaction was not statistically significant, and the small coefficient values (0.005, -0.007, 0.006) showed a very weak influence in predicting outcomes. Hence the interaction terms were not included in the final model.

#### Outliers

Three patients were identified as outliers (Figure 4) which contributed highly influential data points in the logistic regression model that deviated from the expected results (quantified by Cook's distance). Upon data inspection, two of the three patients were found to be presenting with unexpected hospitalisation given their characteristics and symptoms. The two patients were excluded, and the model was re-fitted. The third patient, who presented as an outlier, was not found to have characteristics and symptoms which warranted exclusion from the model; hence this patient was included in the model, despite the high influence, and relatively high Cook's distance (Cooks distance= 0.12, Figure 4).

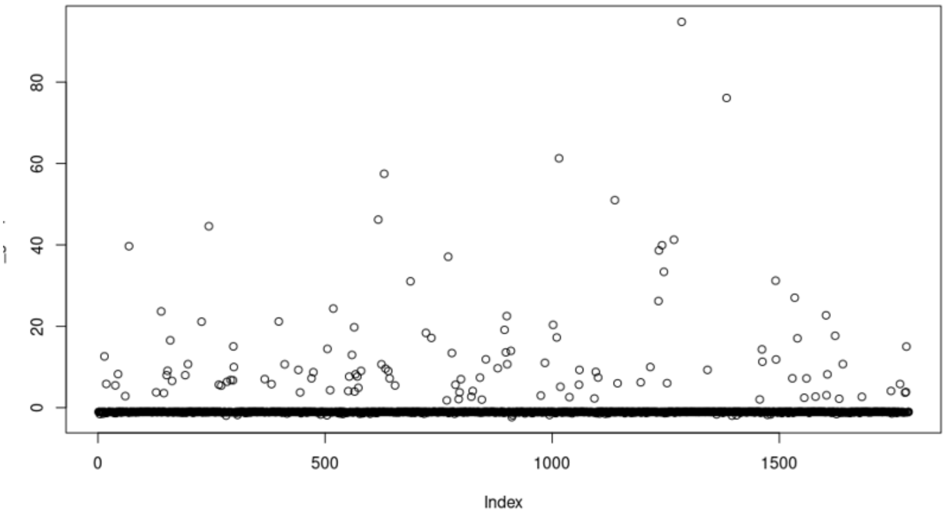


*Figure 4 Plot of residuals for the RCGP-GP model*

As shown in Figure 5 below, the influential outliers were identified by plotting the studentized residuals, representing a standardised residual divided by an estimate of its standard deviation, against the hat values representing the leverage of the patient observation on the logistic regression. By assessing which patients presented a high deviation and high leverage, along with calculating the Cook's distance of each observation to quantify the influence of the observation on the fitted response values, we were able to identify the points that cause the logistic regression to skew towards these outliers and remove them from the final analysis, as necessary.

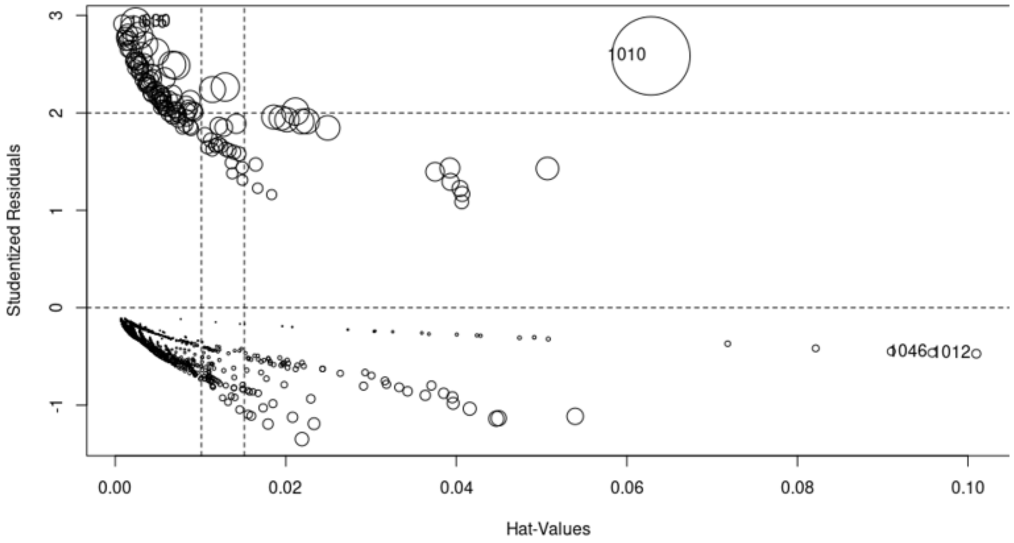


*Figure 5 Plot of studentized residuals and hat values for all patients included in the final model*

#### Internal validation

As part of model internal validation, after bootstrapping, the distributions of the calibration scores in the individually bootstrapped datasets were assessed for normality (Figure 6). These showed that the bootstrapped data generated consistent model results and model diagnostic measures.

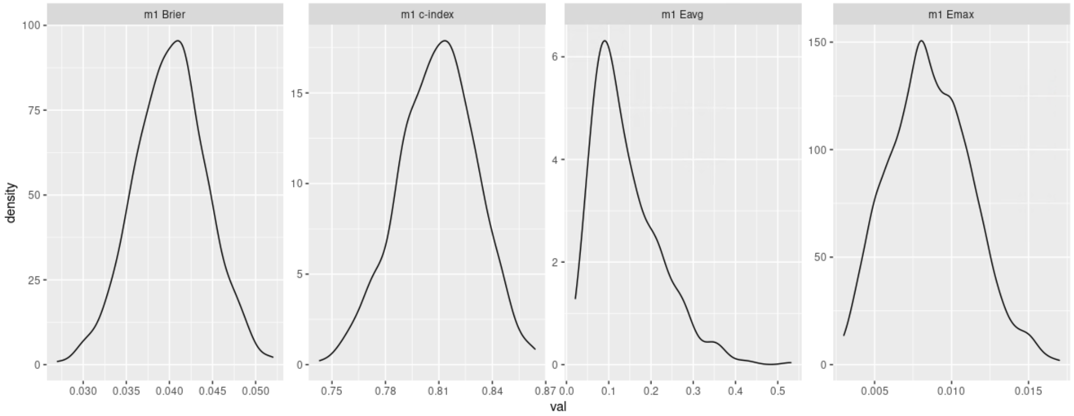


*Figure 6 Distributions of calibration scores in bootstrapped RECAP-GP model*

#### Discrimination analysis

To assess model discrimination, the dataset was split into subpopulations based on sex and age and the final model was validated on the bootstrapped subpopulations datasets. The ROC curves are shown in Figures 7 and Figure 8. The error bars of the models overlapped in all cases, thus there is no significant difference in the predictive power of the model between the defined subpopulations.

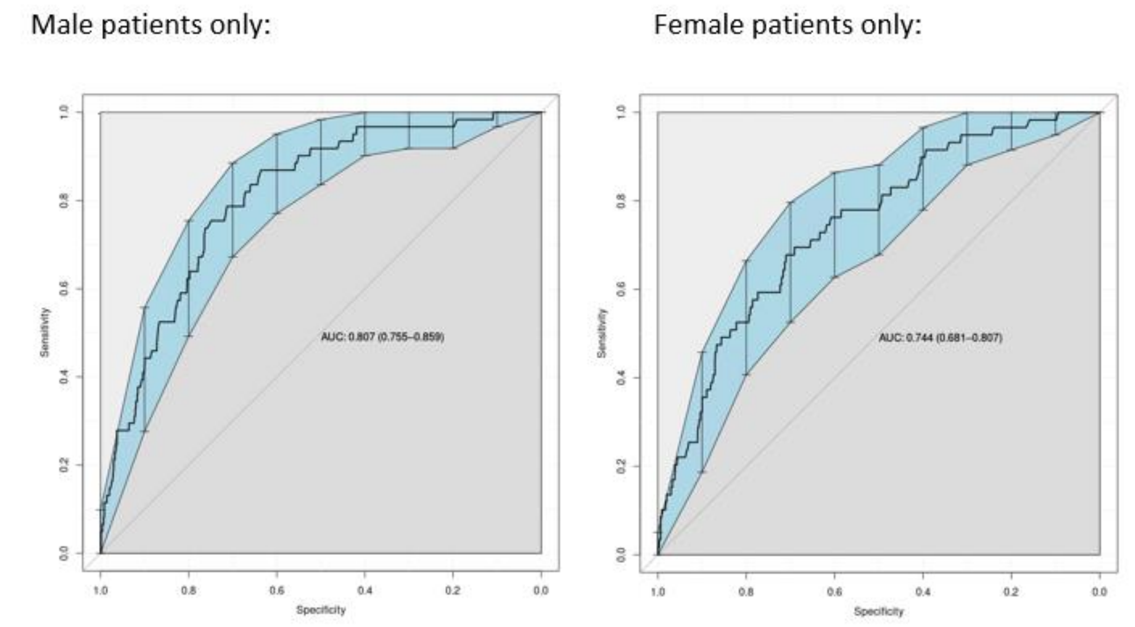


*Figure 7 Gender discrimination analysis of RECAP-GP performance in bootstrapped internal validation dataset*

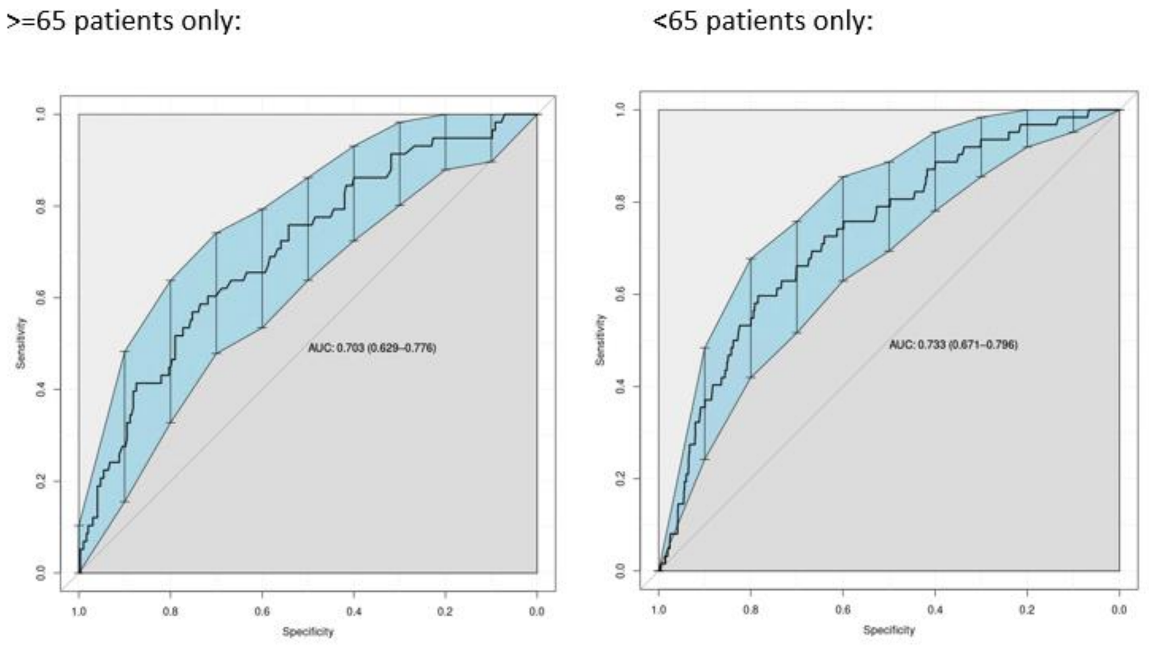


*Figure 8 Age discrimination analysis of RECP-GP performance in bootstrapped internal validation dataset*

#### External validation

The model was externally validated on a dataset from NWL practices and CCAS. The ROC curves are shown in Figures 9 and 10 respectively.

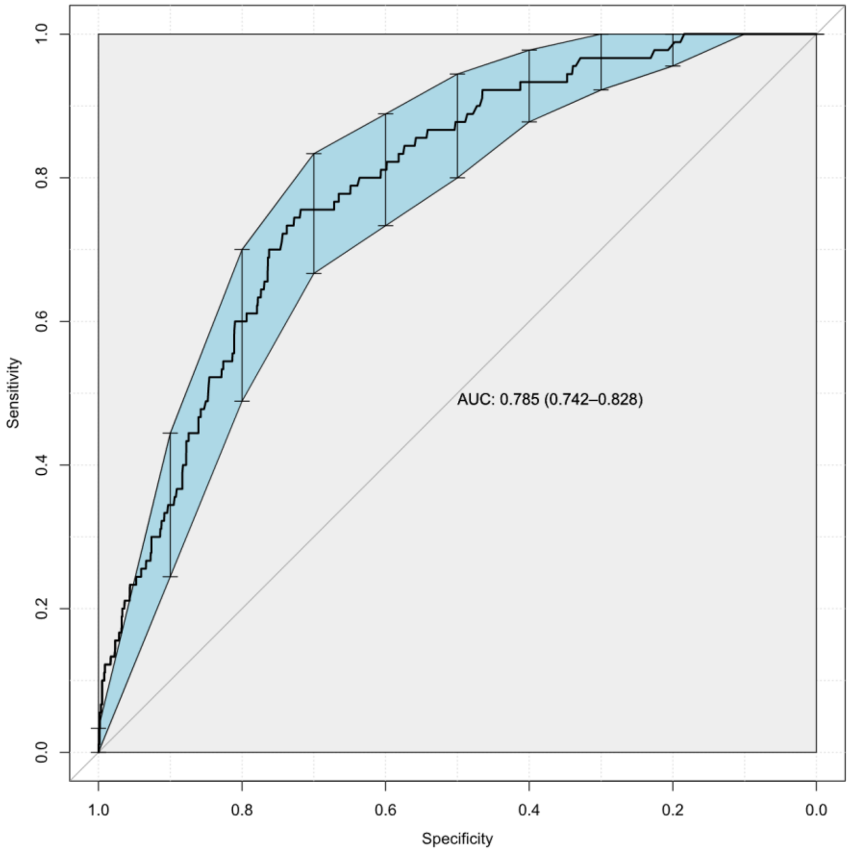


*Figure 9 ROC curve of external validation of the RECAP-GP model on the bootstrapped NWL practice dataset*

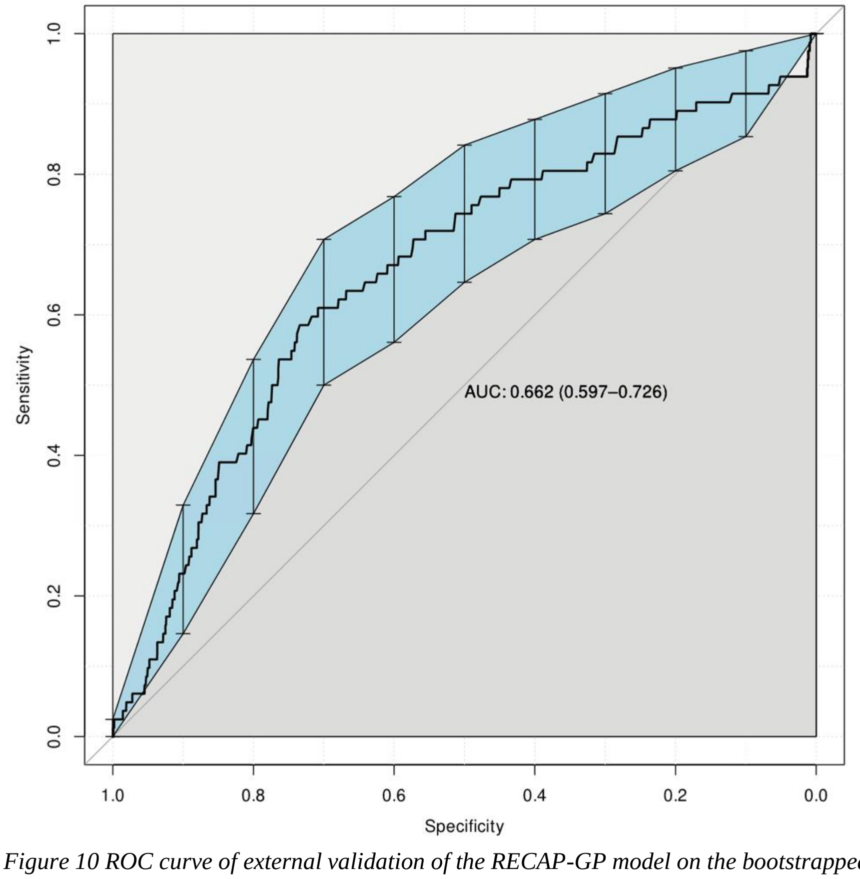


*Figure 10 ROC curve of external validation of the RECAP-GP model on the bootstrapped CCAS practice dataset*

#### RECAP-O2

Figures 11, and Figure 12 below show the age distribution in Doctaly-1 and Doctaly-2 cohorts respectively that were used to develop and validate RECAP-O2 model.

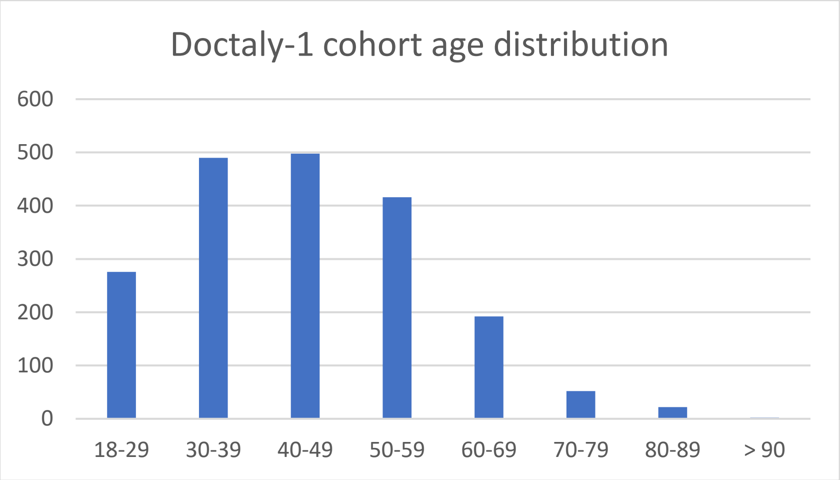


*Figure 11 Age distribution in Doctaly-1 cohort*

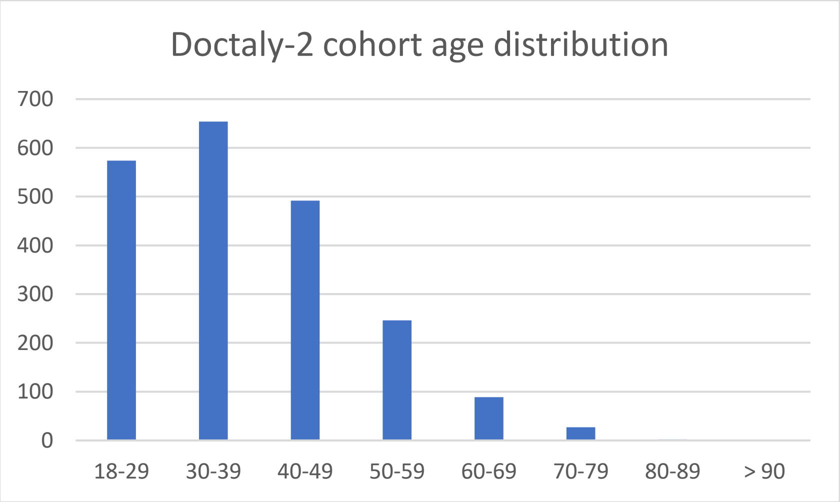


*Figure 2 Age distribution in Doctaly-2 cohort*

##### Parameter non-linearity

The non-linearity of continuous parameters (age and oxygen saturation at rest) was tested using complex splines fitted through penalised thin plane regression. There was no non-linearity in age, and slight non-linearity found in oxygen saturation at rest, estimated as 1.7 degrees of freedom. The degree of non-linearity was assessed by varying the number of the basis dimension used for spline fitting and its effects on the effective number of dimensions in the final fit, with no significant change found. The addition of oxygen saturation at rest as a non-linear term in the model was investigated and no significant improvement in performance was found, with a negligible improvement in AIC of 0.15, and no change in Mc Faddens pseudo R2. Hence, oxygen saturation at rest was included as a linear term as this will be straightforward to implement in the Electronic Health Record (EHR) systems. For age, the dimensionality was optimal at 1 degree of freedom, indicating a linear slope of fit (Figure 13).

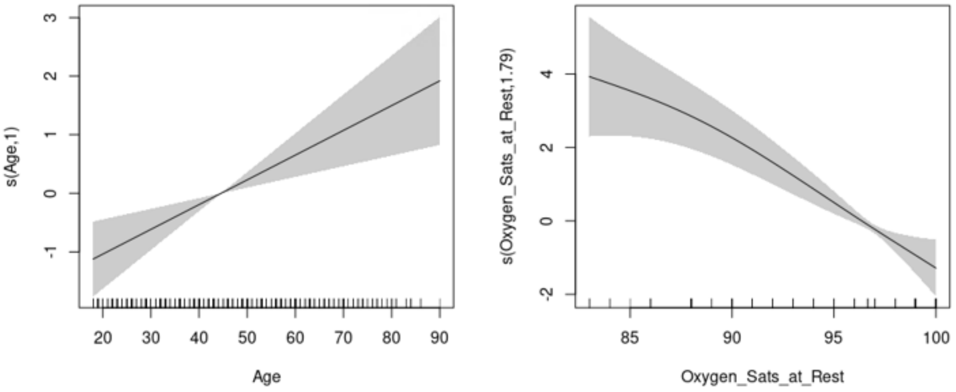


*Figure 13 RCGP-O2 linearity assessment*

#### Outliers

Two patients were identified by visual inspection of a plot of the residuals for each patient prediction (Figure 14 and 15) as outliers contributing highly influential data points that deviated from the expected results (quantified by Cook's distance). Upon data inspection, these two patients were found to be presenting with clinically unexpected hospitalisation given their characteristics and symptoms. Data for these two patients were excluded and the model was re-fitted.

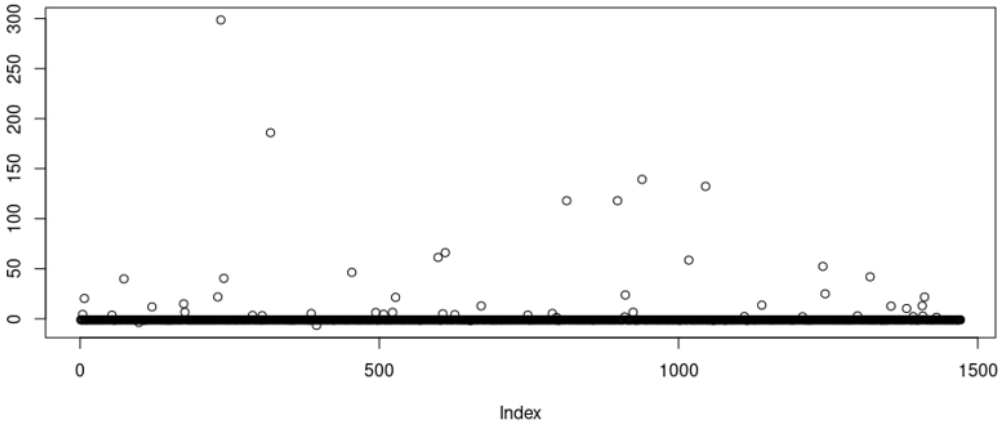


*Figure 14 Plot of residuals for the RECAP-O2 model*

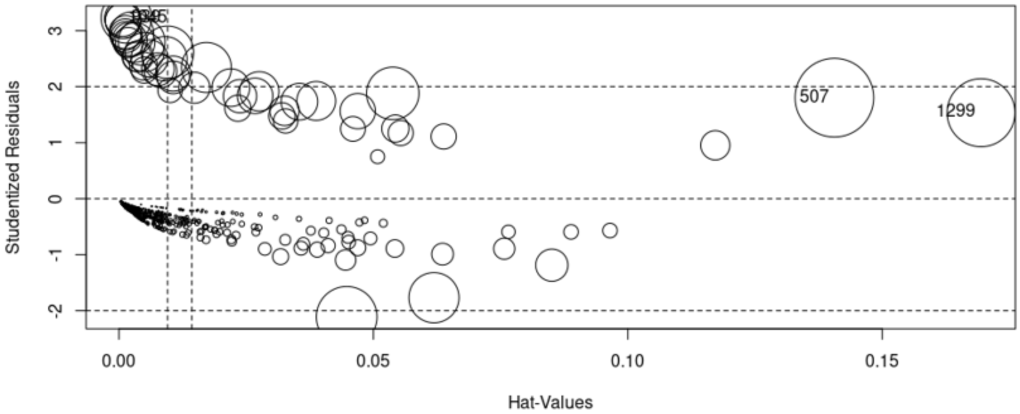


*Figure 15 Plot of studentized residuals and hat values for all patients in the initial model*

#### Oxygen saturation data missingness

The degree of missingness of data in Doctaly was further assessed by inspecting the amount of missing parameter data in time. We found that the first day of recording had the highest level of overall data missingness, especially for blood oxygen saturation measurements, which were the focus for this model. This was due to patients not having immediate access to a pulse oximeter at the time of the first day of recording, but as the SE London Oxygen @home service supplied pulse oximeters, the amount of missing data for oxygen saturation is reduced substantially by the second visit (Figure 16 and Figure 17). Patients with data on only one day of recording were excluded from the analysis, and the final model was developed using data from the second appointment for each patient where available.

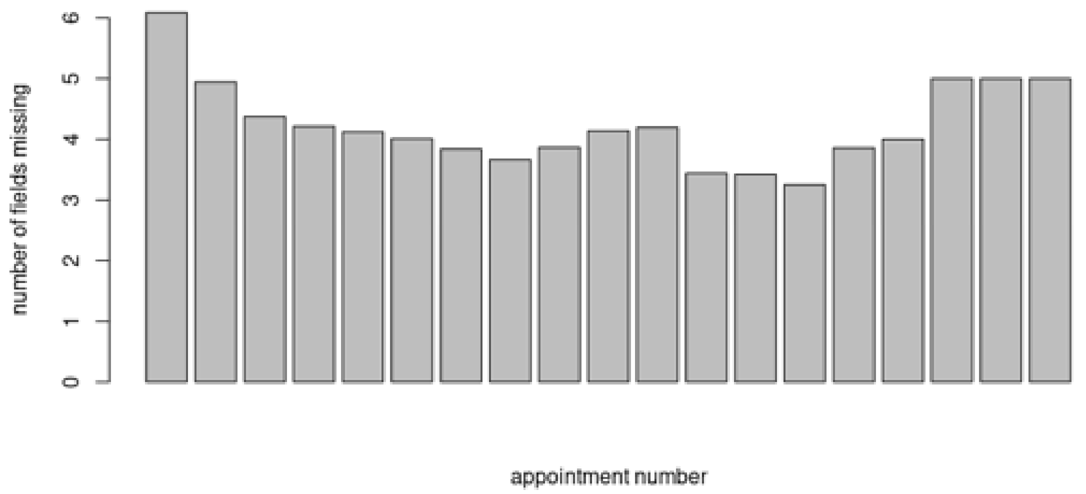


*Figure 16 Mean degree of data missingness for Doctaly-1 patients over each subsequent appointment*

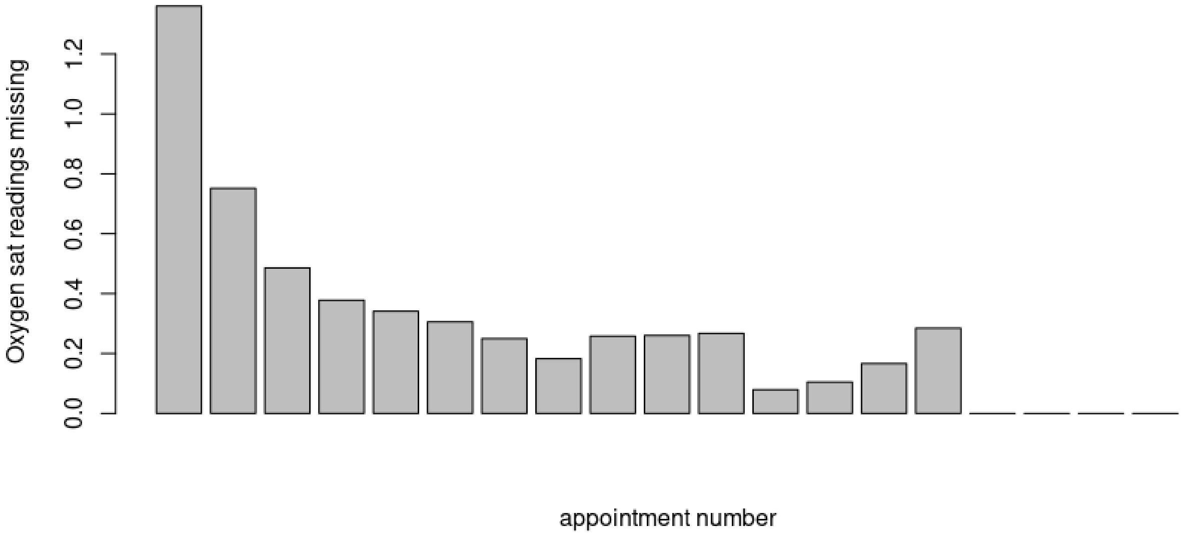


*Figure 10 Mean degree of missingness for oxygen saturation at rest and oxygen saturation after 40 steps readings for Doctaly-1 patients over each subsequent appointment*

#### Internal validation

As part of model internal validation, after bootstrapping, the distributions of the calibration scores in the individually bootstrapped datasets were assessed for normality (Figure 18). These showed that the bootstrapped data generated consistent model results and model diagnostic measures.

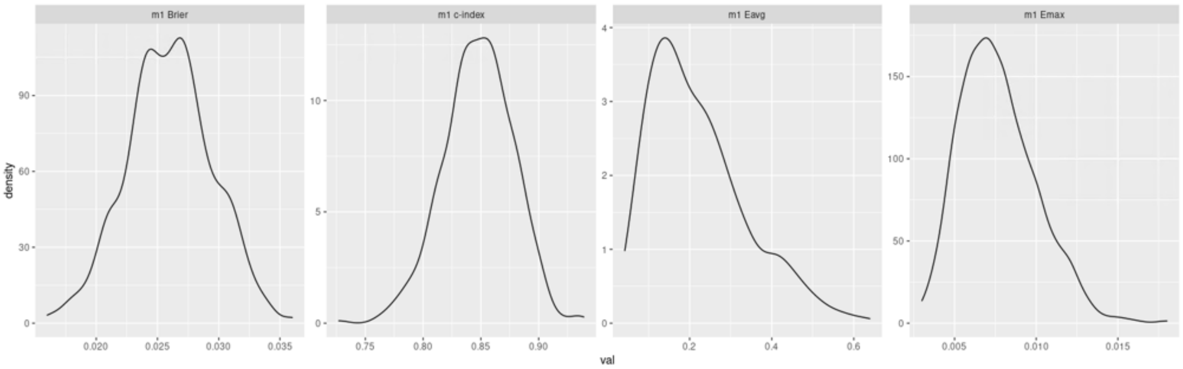

*Figure 18 Distributions of calibration scores in bootstrapped RECAP-O2 model*

#### Discrimination analysis

The discrimination of the model was inspected by splitting the dataset into subpopulations based on sex and age and the final model was validated on the bootstrapped datasets, producing the ROC curves shown in Figure 19 and Figure 20. The confidence intervals for the AUC of the ROC curves (for the bootstrapped dataset) overlapped in all cases, thus there is no significant difference in the predictive power of the model between the defined subpopulations.

The large error bars around the ROC data points in the >=65 age group are due to the small sample and the very low number of hospitalisation events in this age group (Figure 19).

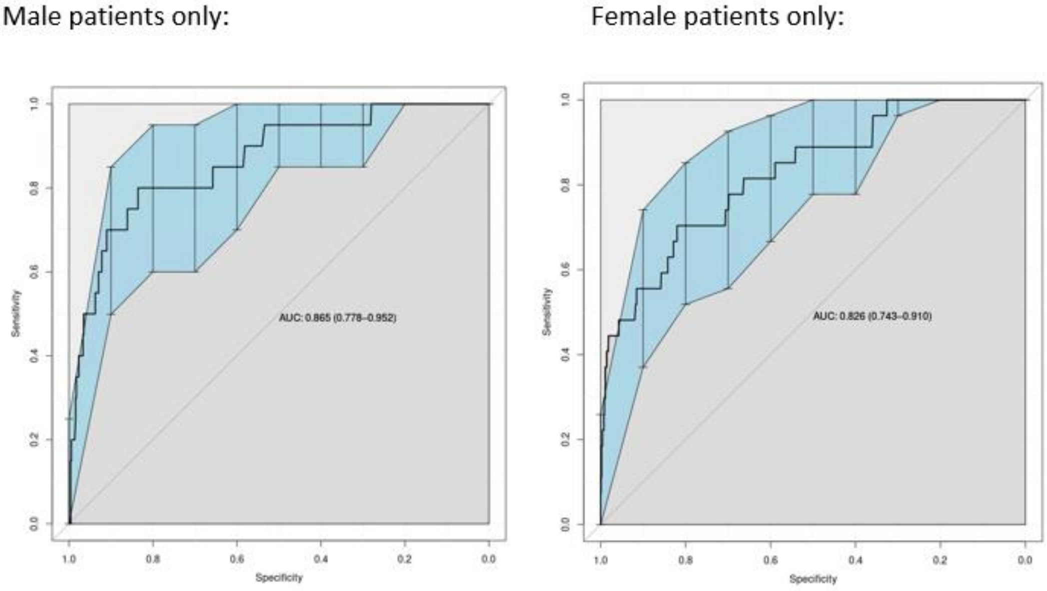

*Figure 19 Gender discrimination analysis of RECAP-O2 performance in bootstrapped internal validation dataset*

>=65 patients only: <65 patients only:

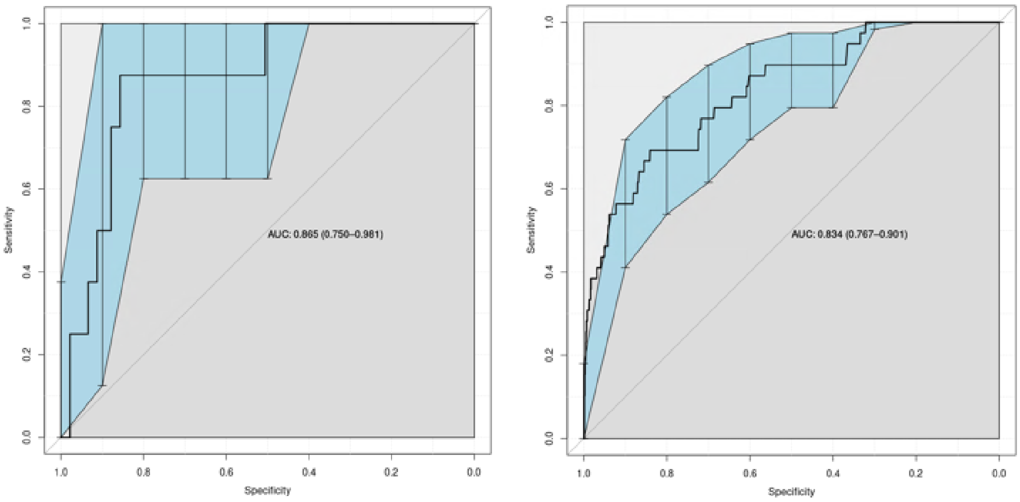


*Figure 20 Age discrimination analysis of RECAP-O2 performance in bootstrapped internal validation dataset*

#### External validation

The model developed on the Doctaly-1 data was externally validated on the Doctaly-2 dataset, and the ROC curve shown in figure NNN was produced. The ROC curve (Figure 21) presents with large error bars at each point and a low AUC because, there are only 19 hospitalisations over 1784 patients, hence a hospitalisation rate of 1%.

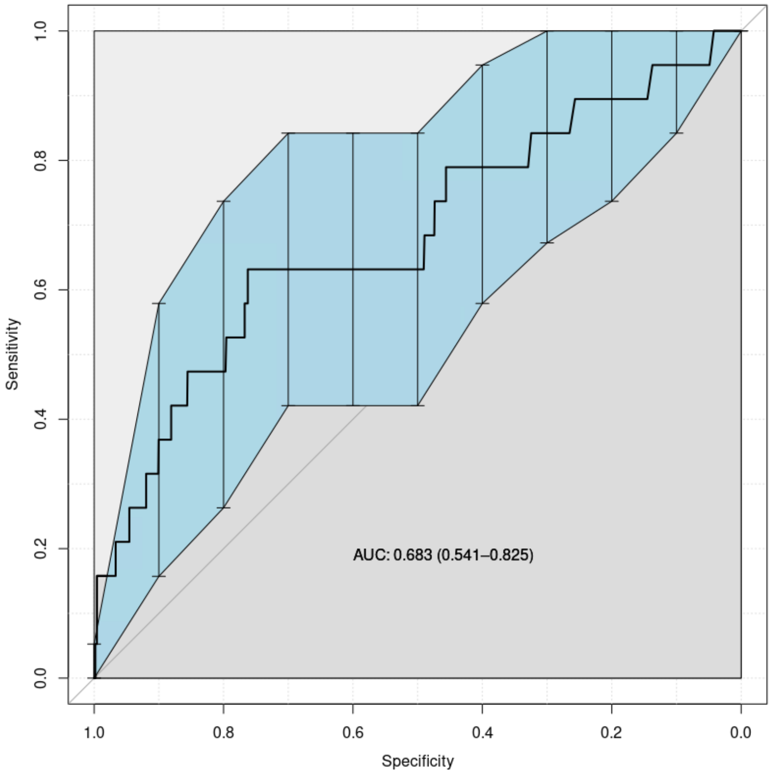


*Figure 21 ROC curve of external validation of the RECAP-O2 model on the bootstrapped Doctaly-2 dataset*

### Doctaly Assist
### NHS Community Coronavirus Monitoring Service
### COVID-19

#### Patient Setup Process

- This will happen at the point the patient is being referred into the COVID-19 Home Monitoring service. The data will be collected via a web-based smart form.
- This data will be the total information the Bot has to make decisions on the flow of patient questions etc. - so important it covers everything that might be needed. We can add additional fields later if required.

#### Data Fields to be captured

1. First Name
2. Surname
3. **NHS Number (Format will be validated)**
4. DOB (From which age will be derived)
5. Phone Number (Mobile Preferred – Format will be validated)
6. Service Referred From: (Drop down: Usual GP, Named Hospital, Named COVID-19 Hot Hub)
7. GP Practice patient is registered to (drop down)
8. Is Patient in “Extremely Vulnerable” Group? YES/NO
9. Is Patient in “High Risk” Group? YES/NO
10. If “YES” to either 8 or 9, please state underlying health reasons (text)
11. **Does patient have an Oxygen Sats Probe? YES/NO**
12. **Does patient have a BP Monitor? YES/NO**
13. **Does patient have an accurate thermometer? YES/NO**
14. **Date when symptoms started?**
15. **Frequency of Remote Clinical Assessment? (Radio Button – 12hrs, 24 hours, 48 hours, 72 hours)**
16. Does patient use WhatsApp and gives consent to be contacted via WhatsApp?
17. **Patient Consent (Y/N) – Gathered during Onboarding Process**

##### “Extremely Vulnerable” Underlying Health Issues – Q8

- Post-transplant
- Active chemo/radiotherapy cancer patients/immunotherapy
- Haematological cancers (at any stage of treatment)
- Severe chest conditions (CF, asthmatics/COPD)
- Rare diseases and inborn errors of metabolism that increase risk of infections including homozygous sickle cell disease
- On immunosuppressant therapy sufficient to increase risk of infection
- Pregnant with heart disease

##### “High Risk” Underlying Health Issue – Q9

- Pregnant
- Over 70s, regardless of medical conditions
- Any adult who qualifies for a flu jab
- Chronic heart disease; kidney; liver disease
- Chronic neurological conditions or learning disability
- Diabetes
- Splenectomy/sickle cell disease
- Weakened immune system: HIV/AIDS, long-term steroids/DMARDs/biologics, having chemotherapy
- Severe obesity (BMI≥40)

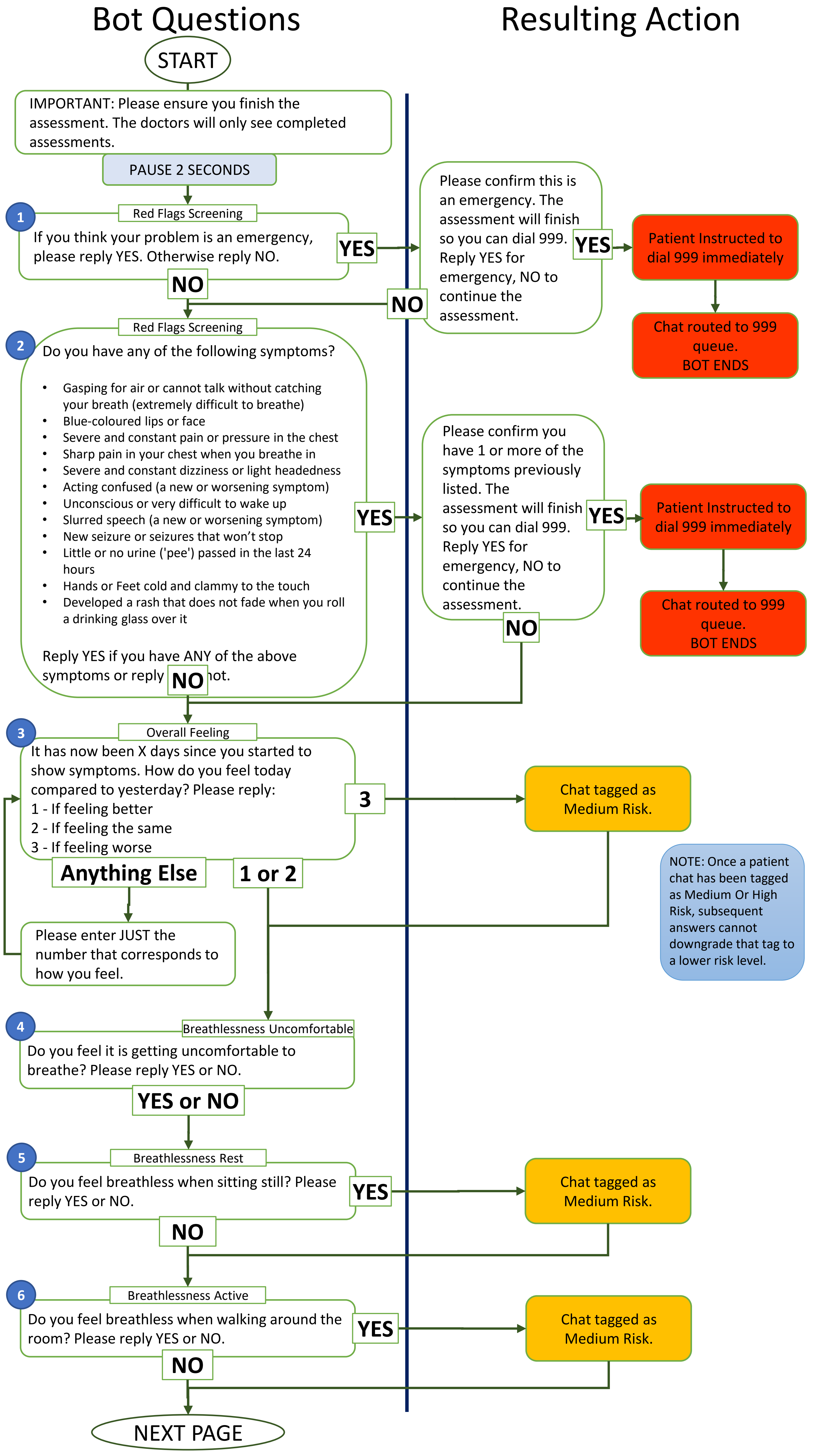

Bot Questions
Resulting Action
START
IMPORTANT: Please ensure you finish the assessment. The doctors will only see completed assessments.
PAUSE 2 SECONDS
1
Red Flags Screening
If you think your problem is an emergency, please reply YES. Otherwise reply NO.
YES
NO
Please confirm this is an emergency. The assessment will finish so you can dial 999. Reply YES for emergency, NO to continue the assessment.
YES
NO
Patient Instructed to dial 999 immediately
Chat routed to 999 queue. BOT ENDS
2
Red Flags Screening
Do you have any of the following symptoms?
• Gasping for air or cannot talk without catching your breath (extremely difficult to breathe)
• Blue-coloured lips or face
• Severe and constant pain or pressure in the chest
• Sharp pain in your chest when you breathe in
• Severe and constant dizziness or light headedness
• Acting confused (a new or worsening symptom)
• Unconscious or very difficult to wake up
• Slurred speech (a new or worsening symptom)
• New seizure or seizures that won't stop
• Little or no urine ('pee') passed in the last 24 hours
• Hands or Feet cold and clammy to the touch
• Developed a rash that does not fade when you roll a drinking glass over it
Reply YES if you have ANY of the above symptoms or reply NO if not.
YES
NO
Please confirm you have 1 or more of the symptoms previously listed. The assessment will finish so you can dial 999. Reply YES for emergency, NO to continue the assessment.
YES
NO
Patient Instructed to dial 999 immediately
Chat routed to 999 queue. BOT ENDS
3
Overall Feeling
It has now been X days since you started to show symptoms. How do you feel today compared to yesterday? Please reply:
1 - If feeling better
2 - If feeling the same
3 - If feeling worse
3
Chat tagged as Medium Risk.
Anything Else
1 or 2
Please enter JUST the number that corresponds to how you feel.
NOTE: Once a patient chat has been tagged as Medium Or High Risk, subsequent answers cannot downgrade that tag to a lower risk level.
4
Breathlessness Uncomfortable
Do you feel it is getting uncomfortable to breathe? Please reply YES or NO.
YES or NO
5
Breathlessness Rest
Do you feel breathless when sitting still? Please reply YES or NO.
YES
NO
Chat tagged as Medium Risk.
6
Breathlessness Active
Do you feel breathless when walking around the room? Please reply YES or NO.
YES
NO
Chat tagged as Medium Risk.
NEXT PAGE

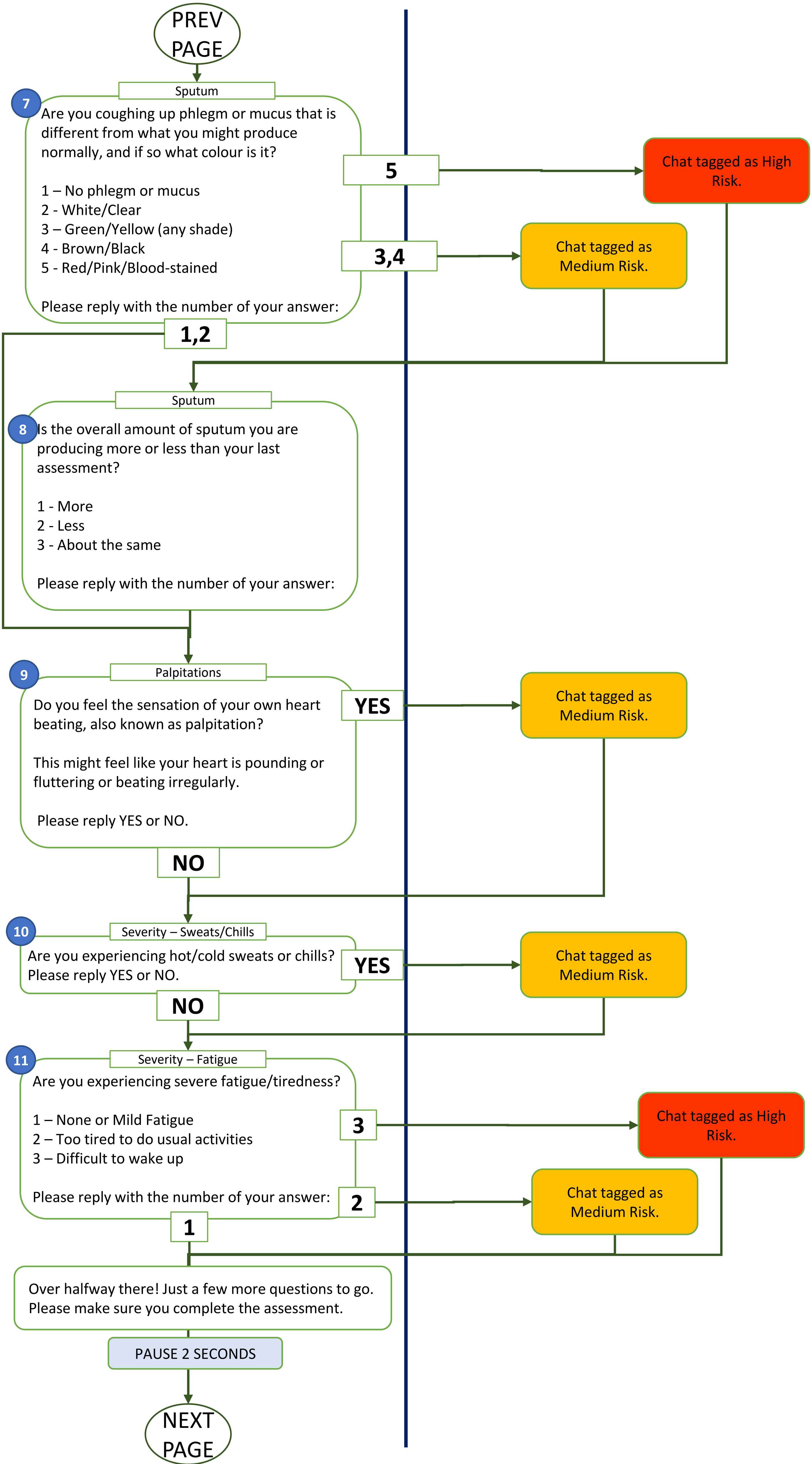
PREV PAGE
7
Sputum
Are you coughing up phlegm or mucus that is different from what you might produce normally, and if so what colour is it?
1 – No phlegm or mucus
2 - White/Clear
3 – Green/Yellow (any shade)
4 - Brown/Black
5 - Red/Pink/Blood-stained
Please reply with the number of your answer:
5
Chat tagged as High Risk.
3,4
Chat tagged as Medium Risk.
1,2
8
Sputum
Is the overall amount of sputum you are producing more or less than your last assessment?
1 - More
2 - Less
3 - About the same
Please reply with the number of your answer:
9
Palpitations
Do you feel the sensation of your own heart beating, also known as palpitation?
This might feel like your heart is pounding or fluttering or beating irregularly.
Please reply YES or NO.
YES
Chat tagged as Medium Risk.
NO
10
Severity – Sweats/Chills
Are you experiencing hot/cold sweats or chills? Please reply YES or NO.
YES
Chat tagged as Medium Risk.
NO
11
Severity – Fatigue
Are you experiencing severe fatigue/tiredness?
1 – None or Mild Fatigue
2 – Too tired to do usual activities
3 – Difficult to wake up
Please reply with the number of your answer:
3
Chat tagged as High Risk.
2
Chat tagged as Medium Risk.
1
Over halfway there! Just a few more questions to go. Please make sure you complete the assessment.
PAUSE 2 SECONDS
NEXT PAGE

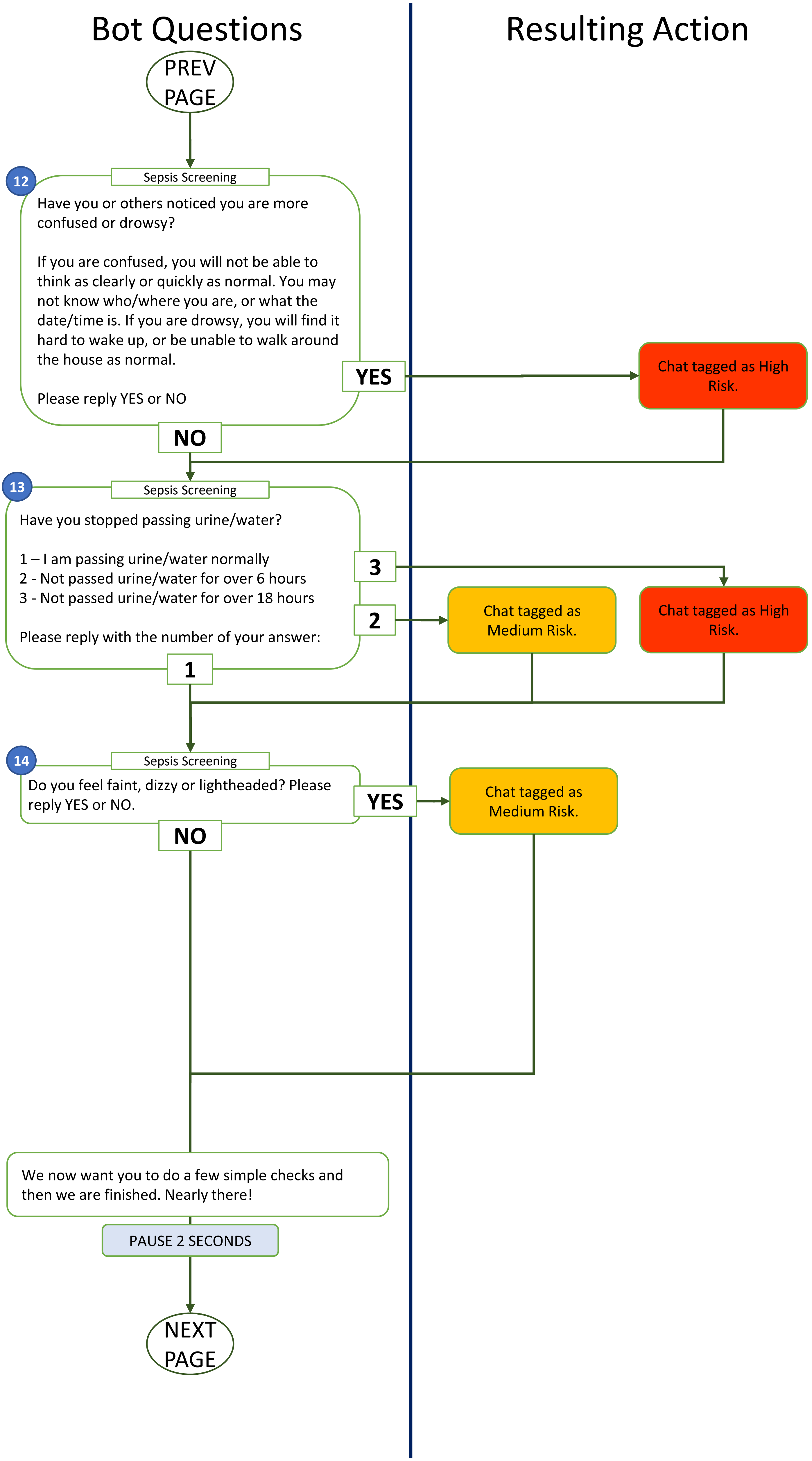
Bot Questions
Resulting Action
PREV PAGE
12
Sepsis Screening
Have you or others noticed you are more confused or drowsy?
If you are confused, you will not be able to think as clearly or quickly as normal. You may not know who/where you are, or what the date/time is. If you are drowsy, you will find it hard to wake up, or be unable to walk around the house as normal.
Please reply YES or NO
YES
Chat tagged as High Risk.
NO
13
Sepsis Screening
Have you stopped passing urine/water?
1 – I am passing urine/water normally
2 - Not passed urine/water for over 6 hours
3 - Not passed urine/water for over 18 hours
Please reply with the number of your answer:
3
Chat tagged as High Risk.
2
Chat tagged as Medium Risk.
1
14
Sepsis Screening
Do you feel faint, dizzy or lightheaded? Please reply YES or NO.
YES
Chat tagged as Medium Risk.
NO
We now want you to do a few simple checks and then we are finished. Nearly there!
PAUSE 2 SECONDS
NEXT PAGE

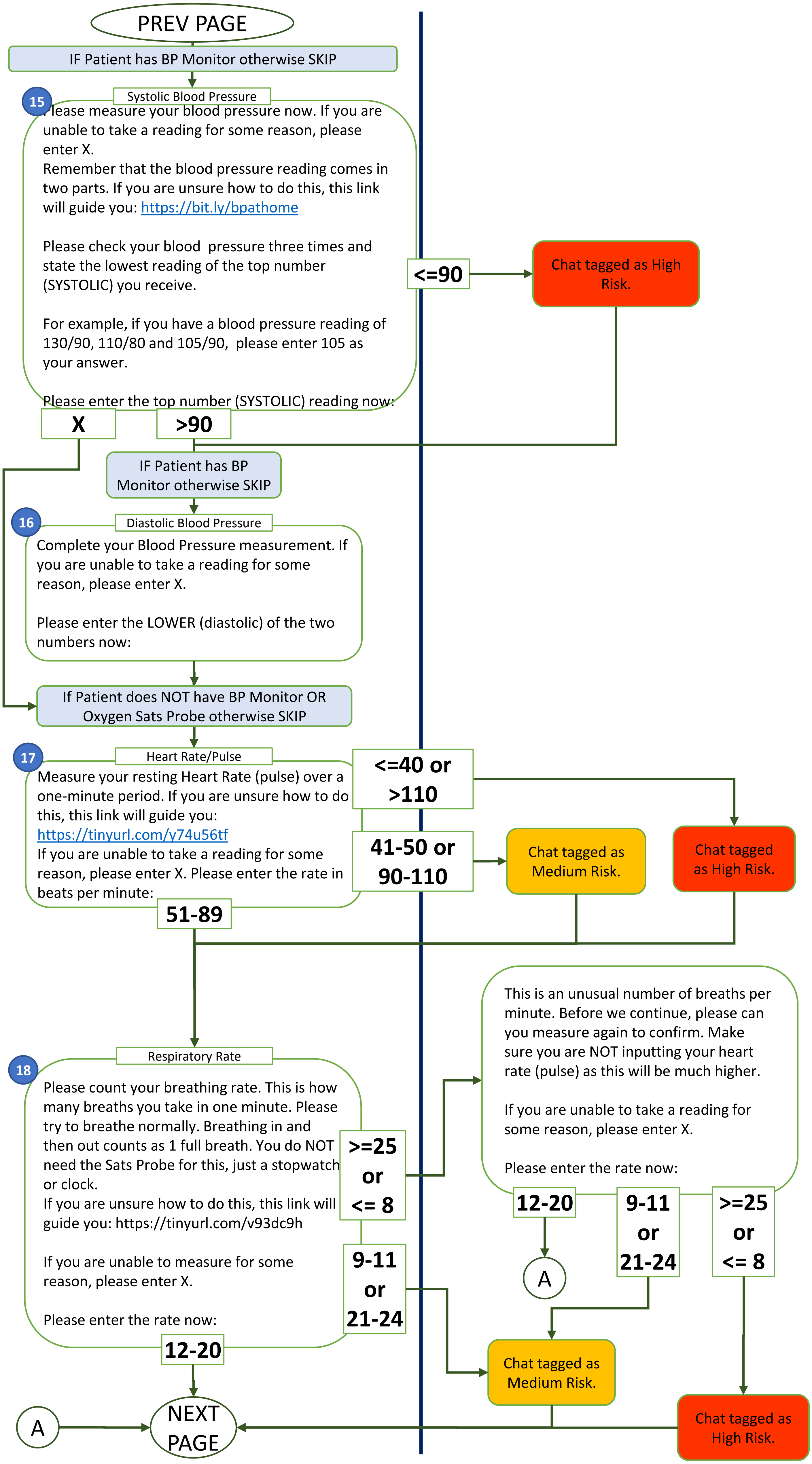
PREV PAGE
IF Patient has BP Monitor otherwise SKIP
15
Systolic Blood Pressure
Please measure your blood pressure now. If you are unable to take a reading for some reason, please enter X.
Remember that the blood pressure reading comes in two parts. If you are unsure how to do this, this link will guide you: https://bit.ly/bpathome
Please check your blood pressure three times and state the lowest reading of the top number (SYSTOLIC) you receive.
For example, if you have a blood pressure reading of 130/90, 110/80 and 105/90, please enter 105 as your answer.
Please enter the top number (SYSTOLIC) reading now:
<=90
Chat tagged as High Risk.
X
>90
IF Patient has BP Monitor otherwise SKIP
16
Diastolic Blood Pressure
Complete your Blood Pressure measurement. If you are unable to take a reading for some reason, please enter X.
Please enter the LOWER (diastolic) of the two numbers now:
If Patient does NOT have BP Monitor OR Oxygen Sats Probe otherwise SKIP
17
Heart Rate/Pulse
Measure your resting Heart Rate (pulse) over a one-minute period. If you are unsure how to do this, this link will guide you: https://tinyurl.com/y74u56tf
If you are unable to take a reading for some reason, please enter X. Please enter the rate in beats per minute:
<=40 or >110
41-50 or 90-110
Chat tagged as Medium Risk.
Chat tagged as High Risk.
51-89
18
Respiratory Rate
Please count your breathing rate. This is how many breaths you take in one minute. Please try to breathe normally. Breathing in and then out counts as 1 full breath. You do NOT need the Sats Probe for this, just a stopwatch or clock.
If you are unsure how to do this, this link will guide you: https://tinyurl.com/v93dc9h
If you are unable to measure for some reason, please enter X.
Please enter the rate now:
>=25 or <= 8
9-11 or 21-24
12-20
This is an unusual number of breaths per minute. Before we continue, please can you measure again to confirm. Make sure you are NOT inputting your heart rate (pulse) as this will be much higher.
If you are unable to take a reading for some reason, please enter X.
Please enter the rate now:
12-20
9-11 or 21-24
>=25 or <= 8
A
Chat tagged as Medium Risk.
Chat tagged as High Risk.
A
NEXT PAGE

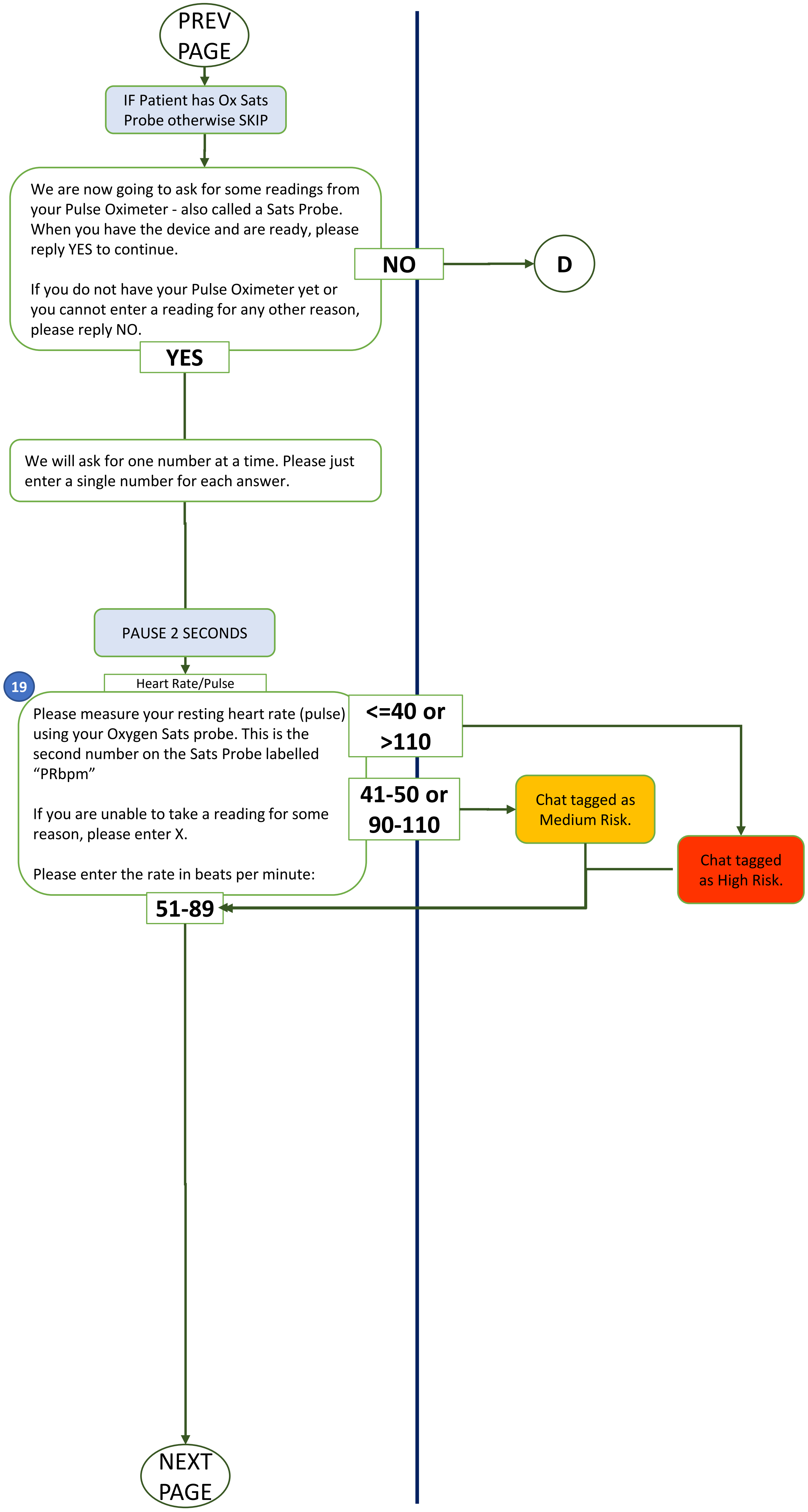
PREV PAGE
IF Patient has Ox Sats Probe otherwise SKIP
We are now going to ask for some readings from your Pulse Oximeter - also called a Sats Probe. When you have the device and are ready, please reply YES to continue.
If you do not have your Pulse Oximeter yet or you cannot enter a reading for any other reason, please reply NO.
NO
D
YES
We will ask for one number at a time. Please just enter a single number for each answer.
PAUSE 2 SECONDS
19
Heart Rate/Pulse
Please measure your resting heart rate (pulse) using your Oxygen Sats probe. This is the second number on the Sats Probe labelled "PRbpm"
If you are unable to take a reading for some reason, please enter X.
Please enter the rate in beats per minute:
<=40 or >110
41-50 or 90-110
Chat tagged as Medium Risk.
Chat tagged as High Risk.
51-89
NEXT PAGE

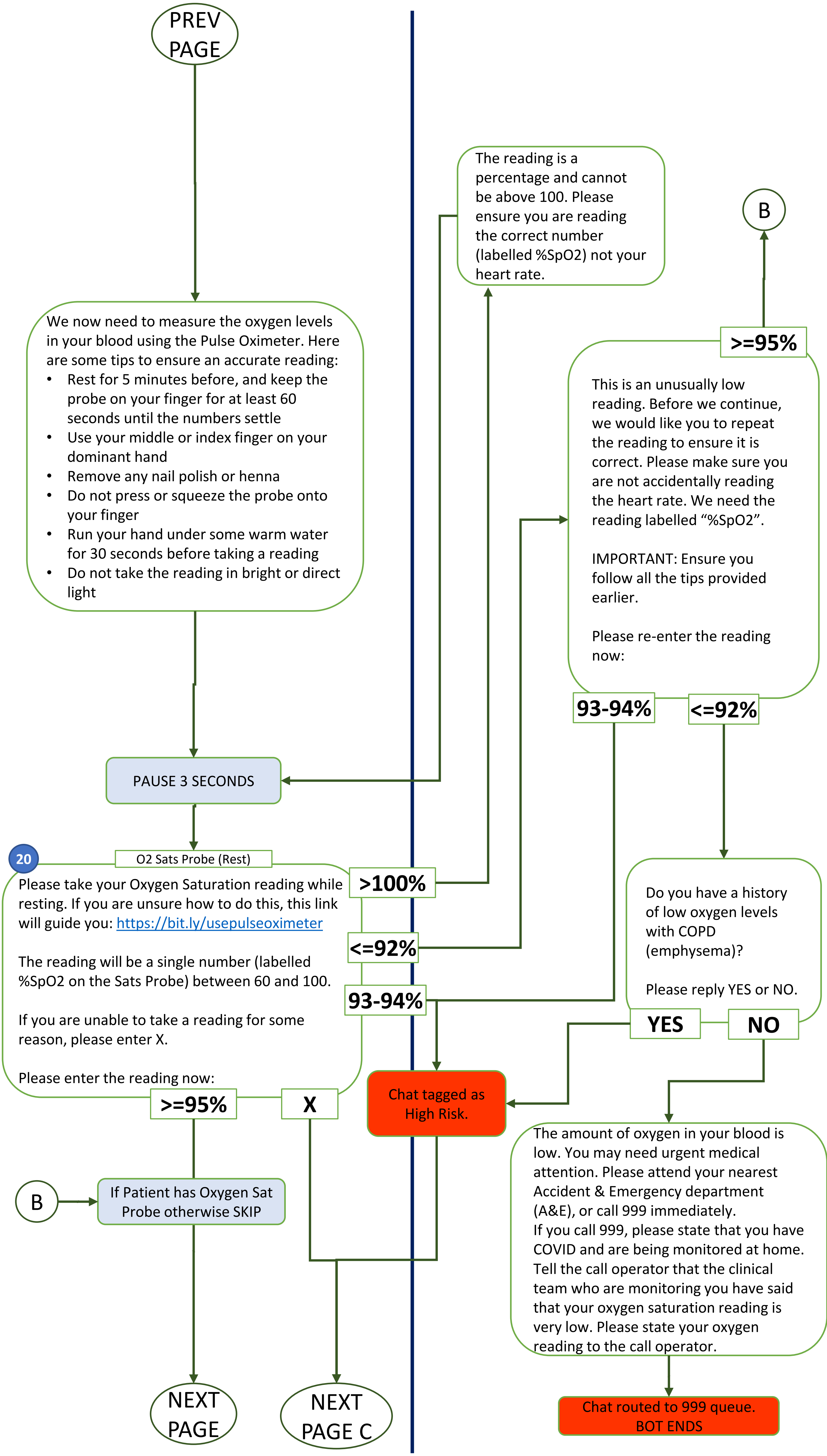
PREV PAGE
We now need to measure the oxygen levels in your blood using the Pulse Oximeter. Here are some tips to ensure an accurate reading:
• Rest for 5 minutes before, and keep the probe on your finger for at least 60 seconds until the numbers settle
• Use your middle or index finger on your dominant hand
• Remove any nail polish or henna
• Do not press or squeeze the probe onto your finger
• Run your hand under some warm water for 30 seconds before taking a reading
• Do not take the reading in bright or direct light
PAUSE 3 SECONDS
20
O2 Sats Probe (Rest)
Please take your Oxygen Saturation reading while resting. If you are unsure how to do this, this link will guide you: https://bit.ly/usepulseoximeter
The reading will be a single number (labelled %SpO2 on the Sats Probe) between 60 and 100.
If you are unable to take a reading for some reason, please enter X.
Please enter the reading now:
>100%
<=92%
93-94%
>=95%
X
The reading is a percentage and cannot be above 100. Please ensure you are reading the correct number (labelled %SpO2) not your heart rate.
B
>=95%
This is an unusually low reading. Before we continue, we would like you to repeat the reading to ensure it is correct. Please make sure you are not accidentally reading the heart rate. We need the reading labelled "%SpO2".
IMPORTANT: Ensure you follow all the tips provided earlier.
Please re-enter the reading now:
93-94%
<=92%
Do you have a history of low oxygen levels with COPD (emphysema)?
Please reply YES or NO.
YES
NO
Chat tagged as High Risk.
B
If Patient has Oxygen Sat Probe otherwise SKIP
The amount of oxygen in your blood is low. You may need urgent medical attention. Please attend your nearest Accident & Emergency department (A&E), or call 999 immediately.
If you call 999, please state that you have COVID and are being monitored at home. Tell the call operator that the clinical team who are monitoring you have said that your oxygen saturation reading is very low. Please state your oxygen reading to the call operator.
Chat routed to 999 queue. BOT ENDS
NEXT PAGE
NEXT PAGE C

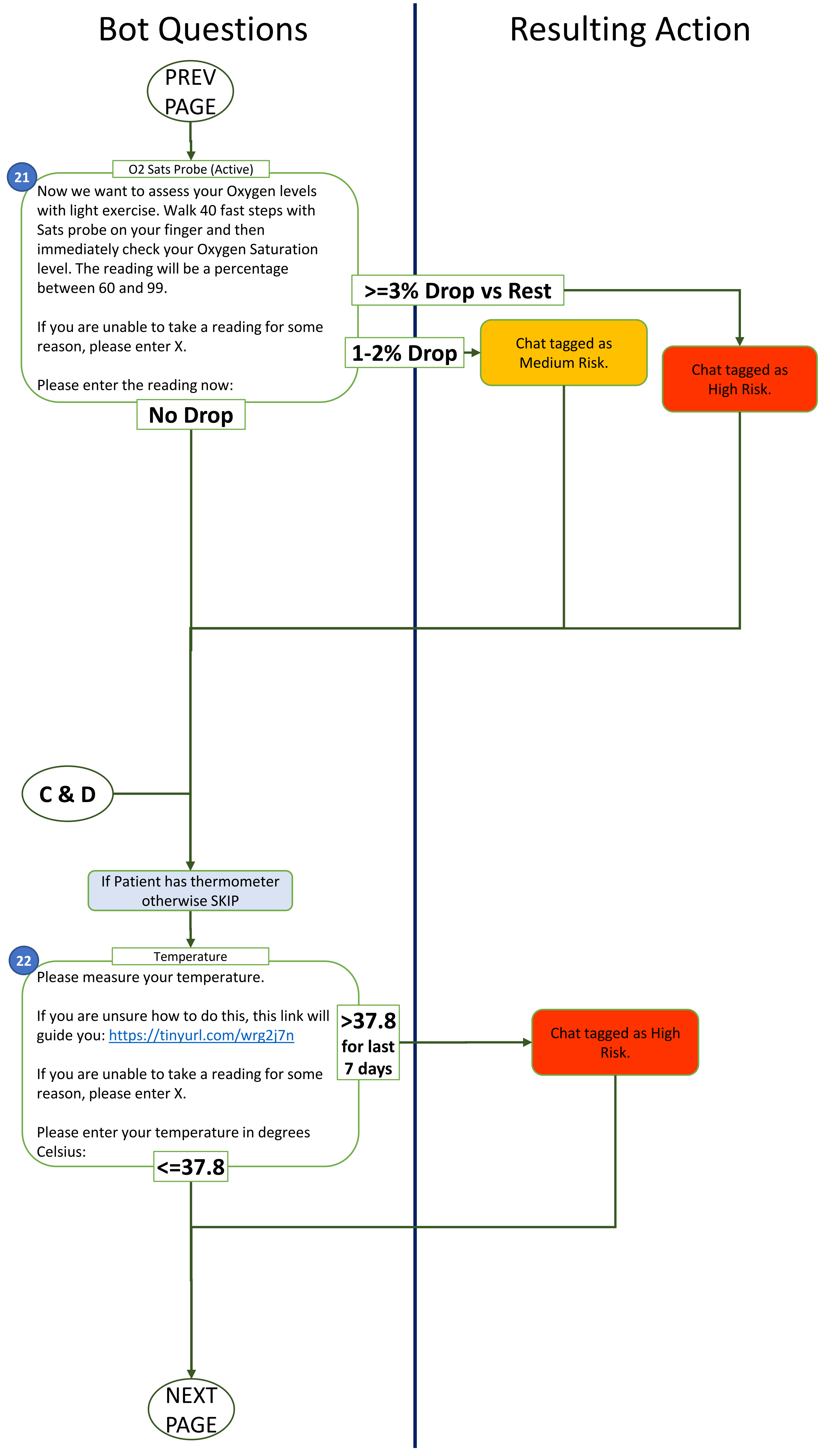
Bot Questions
Resulting Action
PREV PAGE
21
O2 Sats Probe (Active)
Now we want to assess your Oxygen levels with light exercise. Walk 40 fast steps with Sats probe on your finger and then immediately check your Oxygen Saturation level. The reading will be a percentage between 60 and 99.
If you are unable to take a reading for some reason, please enter X.
Please enter the reading now:
>=3% Drop vs Rest
1-2% Drop
Chat tagged as Medium Risk.
Chat tagged as High Risk.
No Drop
C & D
If Patient has thermometer otherwise SKIP
22
Temperature
Please measure your temperature.
If you are unsure how to do this, this link will guide you: https://tinyurl.com/wrg2j7n
If you are unable to take a reading for some reason, please enter X.
Please enter your temperature in degrees Celsius:
>37.8 for last 7 days
Chat tagged as High Risk.
<=37.8
NEXT PAGE

### Bot Questions

### Resulting Action

PREV PAGE

For more information about self-isolation due to COVID-19, please click on these links:

How do I stay safe while isolating?
http://bit.ly/COVIDSafeAtHome

How long do I isolate for?
http://bit.ly/HowLongIsolate

When should I isolate?
http://bit.ly/WhenToIsolate

COMPLETED - SUMMARY

Thank you for completing the assessment on 29/09/2020 03:19PM.
Your responses are logged as follows:

1. Emergency – NO
2. "Red Flag" Symptoms – NO
3. Overall Feeling – BETTER
4. Breathlessness Uncomfortable – NO
5. Breathlessness Rest – NO
6. Breathlessness Active – NO
7. Sputum Colour – BROWN/BLACK
8. Sputum Amount - MORE
9. Palpitations - NO
10. Sweats/Chills – NO
11. Fatigue/Tiredness – LOW
12. Confused or Drowsy – NO
13. Stopped passing urine/water? – NO
14. Faint, dizzy or lightheaded – NO
15. Systolic BP – 120
16. Diastolic BP – 90
17. Manual Heart rate/Pulse – 75
18. Respiratory Rate – 20
19. Heart rate/Pulse – N/A
20. Oxygen Sats at Rest – 99
21. Oxygen Sats Active – 98 (M)
22. Temperature – No Equipment

You have been showing symptoms for 10 days.
You have been using this service for 2 days.
Your current assessment schedule is every 72 hours.

CLINICIAN SUMMARY ACTION: (only if the patient has used the bot for 28 days or more):

- Please note that patient has been using COVI19 chatbot for more than 28 days and so need to consider investigations such as Chest XR or Advice and Guidance to respiratory physician. May need to forward to own GP surgery to action if appropriate.

CLOSE MESSAGE

Your results will now be reviewed by a doctor. If any of the above answers are incorrect and need adjusting, please reply to this message with details.

The doctor may contact you via WhatsApp or phone if any follow up is required. You will be informed of any next steps via WhatsApp including if no action is required.

Is there anything else you would like the doctor to know? They will review any message you reply with when they review your assessment.

If patient answers zero to active Sats confirmation question (COPD) then input Private Note:
"Clinician Note: Patient reports that they suffer from low oxygen levels with COPD; does the patient have type 1 respiratory failure?"

Chat routed to appropriate queue (High, Med, Low)
BOT ENDS
